# Herd immunity induced by COVID-19 vaccination programs and suppression of epidemics caused by the SARS-CoV-2 Delta variant in China

**DOI:** 10.1101/2021.07.23.21261013

**Authors:** Hengcong Liu, Juanjuan Zhang, Jun Cai, Xiaowei Deng, Cheng Peng, Xinghui Chen, Juan Yang, Qianhui Wu, Xinhua Chen, Zhiyuan Chen, Wen Zheng, Cécile Viboud, Wenhong Zhang, Marco Ajelli, Hongjie Yu

**Author notes:** Corresponding authors: Hongjie Yu, Shanghai Institute of Infectious Disease and Biosecurity, School of Public Health, Fudan University, Shanghai 200032, China. These authors contributed equally to this work (Hengcong Liu, Juanjuan Zhang). These authors are joint senior authors contributed equally to this work (Wenhong Zhang, Marco Ajelli, Hongjie Yu).

## Abstract

**Background:** To allow a return to a pre-COVID-19 lifestyle, virtually every country has initiated a vaccination program to mitigate severe disease burden and control transmission. However, it remains to be seen whether herd immunity will be within reach of these programs.

**Methods:** We developed a data-driven model of SARS-CoV-2 transmission for China, a population with low prior immunity from natural infection. The model is calibrated considering COVID-19 natural history and the estimated transmissibility of the Delta variant. Three vaccination programs are tested, including the one currently enacted in China and model-based estimates of the herd immunity level are provided.

**Results:** We found that it is unlike to reach herd immunity for the Delta variant given the relatively low efficacy of the vaccines used in China throughout 2021, the exclusion of underage individuals from the targeted population, and the lack of prior natural immunity. We estimate that, assuming a vaccine efficacy of 90% against the infection, vaccine-induced herd immunity would require a coverage of 93% or higher of the Chinese population. However, even when vaccine-induced herd immunity is not reached, we estimated that vaccination programs can reduce SARS-CoV-2 infections by 53-58% in case of an epidemic starts to unfold in the fall of 2021.

**Conclusions:** Efforts should be taken to increase population’s confidence and willingness to be vaccinated and to guarantee highly efficacious vaccines for a wider age range.

## Introduction

The first-wave of novel coronavirus disease 2019 (COVID-19) in China subsided quickly after the implementation of strict containment measures and travel restrictions starting in March 2020 ^1^. As of August 18, 2021, the COVID-19 pandemic has caused over 208 million reported cases and 4.5 million deaths globally ^2^. The pandemic is far from over, as SARS-CoV-2 has undergone some significant mutations and a number of variants have become widespread due to increased transmissibility and/or immune escape characteristics – e.g., variants Alpha, Beta, Gamma, and Delta. Throughout the globe, a rapid surge of Delta variant cases suggests a clear competitive advantage compared with Alpha, Beta, and Gamma; more than 90% of daily sequences from GISAID are ascribable to the Delta variant since July 2021 ^3^. Despite of no major epidemics, China has been experiencing several minor local outbreaks caused by imported cases of Delta variant, including the outbreaks in Guangzhou, Nanjing, and Zhengzhou city ^1,4^. To suppress transmission, a large share of the world needs to have immunity to SARS-CoV-2, especially to the Delta variant.

Effective vaccines against COVID-19 represent the most viable option to suppress SARS-CoV-2 transmission globally. The effectiveness of vaccination programs depends on several key factors, including vaccine supply, willingness to receive the vaccine, vaccine efficacy, and the age groups targeted by the vaccination effort. However, current vaccination programs are all based on vaccines developed against the historical SARS-CoV-2 lineage, and the efficacy seems be reduced against the Delta variant. In China, home of about 1. billion people (∼18% of the world population), 1.89 billion doses have been administered as of August 12, 2021 ^5^. This figure corresponds to 67.2% of the whole population and 87.6% of the target population (i.e., individuals aged 18 years and above). However, it remains to be seen if and when the vaccine coverage may reach a level sufficient to achieve herd immunity. Countries around the globe are facing the same question.

The classical herd immunity level is defined as 1-1/*R*_*0*_, where *R*_*0*_ is the basic reproduction number – the average number of infections generated by a typical infectious individual in a fully susceptible population ^6^. For a vaccine with efficacy VE that gives life-long protection, the level of herd immunity required to stop transmission is (1-1/*R*_*0*_)/VE. However, this estimate is an oversimplification of a complex phenomenon as it ignores the heterogeneities of actual human population (e.g., social mixing patterns, age-specific susceptibility to infection) ^7,8^ as well as of vaccination (e.g., lifelong immunity, sterilizing vaccine). To overcome this limitation, here we integrate contact survey specific of the Chinese population ^9^ as well as official demographic statistics to develop an age-structured stochastic model to simulate SARS-CoV-2 transmission (Fig.S1 in *SI Appendix*). We then use this model to evaluate whether herd immunity is achievable against the Delta variant or not via mass vaccination and to explore the way forward to achieve suppression of transmission.

## Methods

### SARS-CoV-2 transmission and vaccination model

We built a compartmental model of SRAS-CoV-2 transmission and vaccination, based on an age-structured stochastic SIR scheme, accounting for heterogeneous contact patterns by age ^9^ and heterogeneous susceptibility to infection by age as estimated using contact tracing data in Hunan province of China. In the model, the population is divided into three epidemiological categories: susceptible, infectious, and removed, stratified by 17 age groups. Susceptible individuals can become infectious after contact with an infectious individual according to the age-specific force of infection. The rate at which contacts occur is determined by the mixing patterns of each age group. The average generation time was set to 7 days. We consider a basic reproductive number (*R*_*0*_) of 6.0 according to estimates for the SARS-CoV-2 Delta variant. Simulations are initiated with 40 infectious individuals, corresponding to the number of cases first detected in a local outbreak in Beijing on June 11, 2020. We consider a 2-dose vaccine, that only susceptible individuals are eligible for vaccination (we recall that natural immunity is close to 0 in China as of July 2021), and that the duration of vaccine-induced immunity lasts longer than the time horizon considered in this study (i.e., 1 year). Details about the model and parameters are reported in Sec. 1 of *SI Appendix*.

The model allows the explicit simulation of the vaccination strategy currently used in China: random distribution of vaccines to adults aged 18+ years (strategy 1). Two of the SARS-CoV-2 vaccines (i.e., BBIBP-Corv and CoronaVac) used in China have been licensed for children aged 3 to 17 years ^10^; however, as of August 12, 2021 children are not included among the target population for vaccination in most provinces. To explore the contribution of vaccinating children aged 3-17 years to achieving herd immunity, we test two alternative strategies: i) same as strategy 1, but extended to individuals aged 3+ years starting from September 1, 2021 (strategy 2); ii) random distribution of vaccines to individuals aged 3+ years since the start of vaccination programs (strategy 3).

### Baseline scenario

As the baseline scenario, we considered the following assumptions:

i. **Epidemic seeding**: An epidemic is assumed to be triggered by forty SARS-CoV-2 infections on September 1, 2021; vaccines have been rolling out in China since November 30, 2020.
ii. **Vaccination strategy:** We test three different vaccination strategies:
  a. Strategy 1—random distribution of vaccines to adults aged 18+ years
  b. Strategy 2—same as strategy 1, but the vaccination is extended to individuals aged 3+ years starting from September 1, 2021
  c. Strategy 3—random distribution of vaccines to individuals aged 3+ years since the start of the vaccination program. Note that in all scenarios, we consider that a fraction of the population (about 2% - Sec. 4 of *SI Appendix*) is not eligible to receive the vaccine (e.g., pregnant women, individuals with allergies or other conditions preventing them to safely receive the vaccine).
iii. **Vaccine capacity**: We simulated the daily vaccine administration capacity based on the vaccine capacity data throughout the entire vaccination campaign in China ^5^. We found that the daily vaccine administration capacity exponentially increased during the initial phase of the campaign before stabilizing at 294,234 doses per day on June 2, 2021 for the Shanghai population (about 24 million individuals; details are reported in Sec. 5 of *SI Appendix*).
iv. **Vaccine efficacy (VE)**: The vaccine schedule requires two doses and VE is estimated at 54.3% against the infection for the Delta variant. This estimate is based on the efficacy measured against the historical lineages and the reduction of neutralizing antibodies estimated for Delta variant in clinical studies (see Tab. S1 of *SI Appendix* for details).We also explore higher VE values as sensitivity analyses (Tab. S1 of *SI Appendix*). We also test a two-dose schedule with a 14-day interval. In addition, COVID-19 vaccines may not be equally effective across age groups in preventing infection. To understand the impact of this assumption, we also tested a relative VE of 50% and 75% for individuals aged 3-17 and 60+ years as compared to VE for individuals aged 18-59 years.
v. **Vaccine action**: We considered two ways in which VE could be below 100%: an “all-or-nothing” vaccine (baseline analysis), in which the vaccine provides full protection to a fraction VE of individuals who are vaccinated and no protection to the remaining 1-VE vaccinated individual. The second option we considered is a “leaky” vaccine in which all vaccinated individuals have a certain level of protection to the infection corresponding to VE.
vi. **Initial immunity**: As of July 2021, there is essentially no population immunity from natural infection in China. For the sake of generalizability of results to other countries with ongoing transmission, we have explored a scenario where 30% of the population has initial natural immunity.
vii. **Susceptibility to infection by age**: Children under 15 years of age were estimated to have a lower susceptibility to SARS-CoV-2 infection as compared to adults (i.e., individuals aged 15 to 64 years), while individuals aged 65+ years had the highest susceptibility to infection.
viii. **Immunity duration**: We let the transmission model run for one year, assuming a life-long protection from natural infection or vaccination.

Comprehensive sensitivity analyses to evaluate the impact of the baseline assumptions on our results are carried out as well (Tab. S1 in *SI Appendix*).

### Alternative vaccination scenarios

We test three alternative scenarios to explore the potential for vaccination-induced herd immunity, where: i) epidemic start is delayed from September 1, 2021 to October 1, 2021, and November 1, 2021; ii) the initial reproduction number varies between 1.1 and 6 to account for different intensity of NPIs 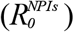; iii) combinations of scenarios i and ii. For scenario ii), we do not explicitly model single non-pharmaceutical interventions (NPIs) such as case isolation, contact tracing, wearing masks, social distancing, improved hygiene. Instead, the synergetic effect of these measures is considered as a reduction of the reproduction number.

### Data analysis

For each scenario, 100 stochastic simulations were performed. The output of these simulations determined the distribution of the number of infections. We defined 95% credible intervals as quantiles 0.025 and 0.975 of the estimated distributions.

We used the next-generation matrix approach to estimate the reproduction number, *R*_*e*_. Herd immunity is considered as achievable when *R*_*e*_ <1. Details are reported in Sec. 2-3 in *SI Appendix*.

## Results

### Baseline scenario

By forward simulating one year of epidemic and assuming no vaccine hesitancy, continued vaccination efforts would lead to a final coverage of 97.6% of the target population, which corresponds to 86.4% of the total population for strategy 1 (Fig. 1A). For strategy 2 and 3, the estimated coverage of the total population increases to 93.0% and 95.0%, respectively (Fig. 1B-C). Under any scenario, the mean incidence of newly infected individuals never reaches 250 over 10,000 residents (Fig. 1D-F). We estimated that the effective reproduction number at the time the infection is seeded (*R*_*e*_) is still well above the epidemic threshold, namely 4.50 (95%CI: 3.80-4.98), 4.49 (95%CI: 3.90-4.89), and 3.67 (95%CI: 3.64-3.68) for strategy 1-3, respectively. These estimates suggest that the vaccine coverage on September 1, 2021 is not enough to prevent onward transmission, regardless of the vaccination strategy. *R*_*e*_ is estimated to cross the epidemic threshold (i.e., 1) on October 15, October 16, and October 21, 2021 for strategy 1-3, respectively, due to the accumulation of immune individuals both through the continue vaccination efforts and natural infections (Fig. 1G-I). The estimated infection attack rates are 47.3% (95%CI: 45.8-48.6%), 44.0% (95%CI: 42.3-45.9%), and 42.3% (95%CI: 39.5-44.0%) for strategy 1-3, respectively (Fig.1J-L).

**Figure 1.**
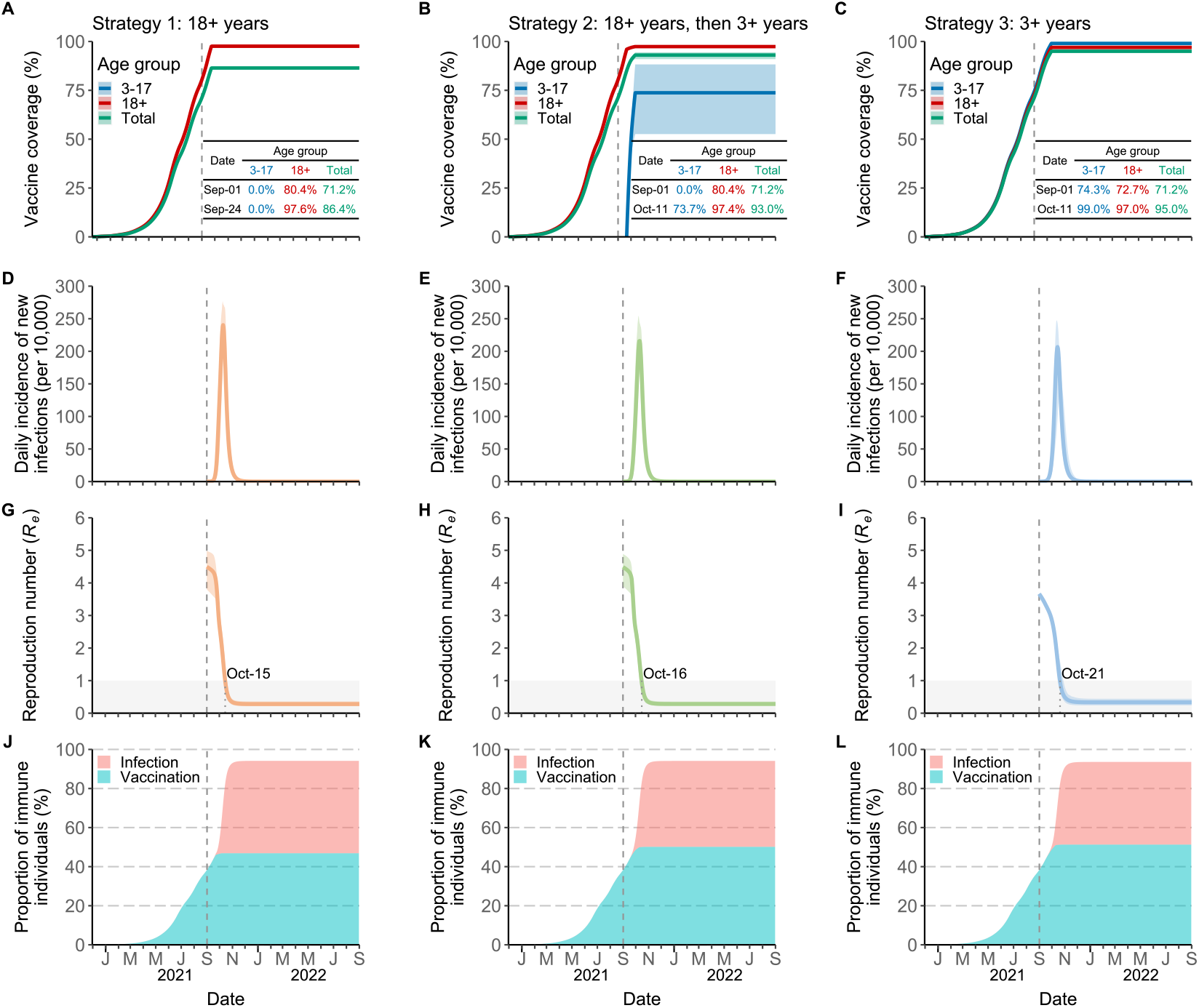
Time series of vaccine coverage, daily incidence of new SARS-CoV-2 Delta variant infections, effective reproductive number, *R*_*e*_, and fraction of immune population. **A** Age-specific vaccine coverage over time for strategy 1. Vaccination program is assumed to be initiated on November 30, 2020 (i.e., the time that the vaccine doses administrated was first officially reported in China). The dotted lines correspond the start of epidemic. The inserted table shows the age-specific coverage for the two key time points (the start of epidemic (i.e., September 1, 2021), and the time that the coverage keeps constant (i.e., September 24), respectively). The line corresponds to the mean value, while the shaded area represents 95% quantile intervals (CI). **B** As A, but for strategy 2. **C** As A, but for strategy 3. **D** Daily incidence of new SARS-CoV-2 infections per 10,000 individuals for strategy 1 (mean and 95% CI). **E** As D, but for strategy 2. **F** As D, but for strategy 3. **G** Effective reproduction number *R*_*e*_ over time (mean and 95% CI), as estimated using the Next-Generation matrix method from the time series of susceptible individuals for strategy 1. The shaded area in gray indicates the epidemic threshold *R*_*e*_ =1. The numbers around the shaded area indicate when *R*_*e*_ cross this threshold (i.e., October 15) for strategy 1. **H** As G, but for strategy 2. **I** As G, but for strategy 3. **J** Proportion of immune population due to either natural infection or vaccination over time for strategy 1. **K** As J, but for strategy 2. **L** As J, but for strategy 3.

Although vaccine-induced immunity is not enough to prevent viral circulation, all the scenarios considered are associated with substantial mitigation of COVID-19 burden. We estimate the infection attack rate for the three vaccination strategies to decrease by more than 50% with respect to a reference scenario with no interventions. Strategy 2-3 achieve the highest reduction (55.3% and 57.0%) thanks to the inclusion of the age group 3-17 years in the target population of the vaccination program (Fig. 2A-B).

**Figure 2.**
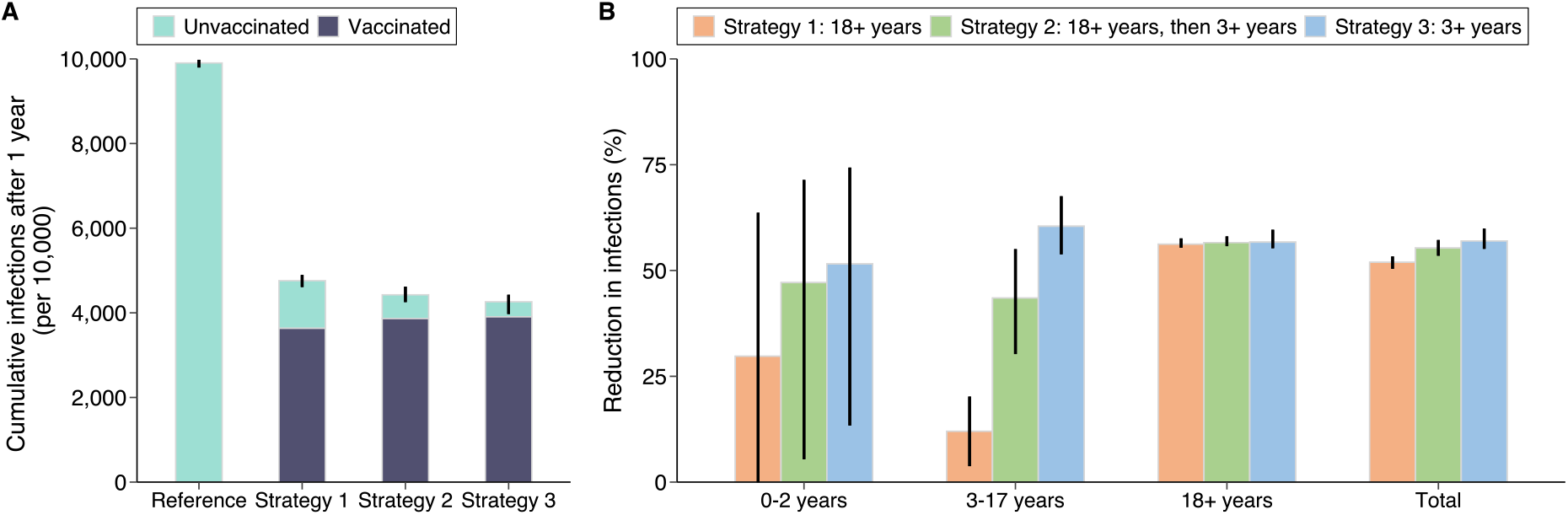
Burden of COVID-19 in the baseline scenario. **A** Cumulative number of infections per 10,000 individuals after 1 simulated year for *reference scenario* and three vaccination strategies (mean and 95% CI). *Reference scenario* indicates no vaccination and no NPIs with *R*_*0*_=6.0 at the beginning of transmission. Infections consist of unvaccinated and vaccinated individuals. The bar corresponds to the mean value, while the vertical line represents 95% quantile intervals. **B** Reduction in infections (mean and 95% CI) with respect to the *reference scenario* in different age groups and the total population. The reduction is defined as the estimated number of infections after 1 year since the introduction of the initial infected individuals under *reference scenario* minus the one under the vaccination strategy, relative to the estimated number under *reference scenario*. The 95% CI of the reduction may cross 0 as the burden between reference scenario and vaccination scenario is approximately the same in some simulations. We thus trimmed the lower limit of 95% CI at 0 through the manuscript.

These results were based on the assumption of an “all-or-nothing” vaccine (i.e., a vaccinated individual will either develop full protection with probability given by the vaccine efficacy or zero protection). To test the robustness of our findings to this assumption, we also tested a “leaky” vaccine (i.e., the susceptibility to infection of any vaccinated individual is reduced by a factor equal to the vaccine efficacy ^11^) and we obtained similar results with respect to the baseline analysis (Fig. S3 in *SI Appendix*). Moreover, the obtained results are confirmed when the initial number of seeds is varied in the range from 10 to 100 (Fig. S4 in *SI Appendix*), and when equal susceptibility to infection by age is assumed (Fig. S5 in *SI Appendix*). Finally, we also conducted a counterfactual analysis where we assume that a part of the population was already immune before the start of the vaccination campaign (similar to the situation in Western countries). Under this assumption, we found that a 30% initial immunity proportion would not lead to *R*_*e*_ below the epidemic threshold for strategy 3 before September 1, the start date of the simulations (Fig. S6 in *SI Appendix*).

As regard the parameters regulating the vaccination process, we found that the (overall) vaccine efficacy has the largest impact, followed by the vaccine efficacy of individuals aged 3-17 and 60+ relative to individuals aged 18-59 years (Fig. S7 and S8 in *SI Appendix*). On the other hand, the time between vaccination and maximum protection and the time interval between the first and second dose have a more moderate impact on the overall effectiveness of the analyzed vaccination strategies (Fig. S9 and S10 in *SI Appendix*).

### Scenario 1: Delaying the start of the epidemic

The findings presented thus far suggest that herd immunity against Delta variant cannot be built through vaccination by September 1, 2021. Next, we test to what extent the start of a new epidemic wave needs to be delayed (e.g., by keeping strict restriction for international travels) to allow the immunity to build up in the population, potentially reaching herd immunity levels. According to the daily vaccine capacity used in the baseline scenario (based on the history of daily vaccination capacity data up to May 23, 2021), we estimate that *R*_*e*_ remains above the epidemic threshold for all three strategies even if the seeding of an epidemic is delayed to November 1, 2021 - before that time, the vaccination coverage has reached the maximum in the target population (i.e., 97.6%, 97.8%, and 97.8% for strategy 1-3) (Fig. 3A and Fig. S11 in *SI Appendix*). In addition, in this case, strategies 2 and 3 lead to a higher reduction of the infection attack rate with respect to the scenario with no intervention (less than 4,200 per 10,000 individuals for an epidemic starting on November 1, 2021 - Fig. 3B-C).

**Figure 3.**
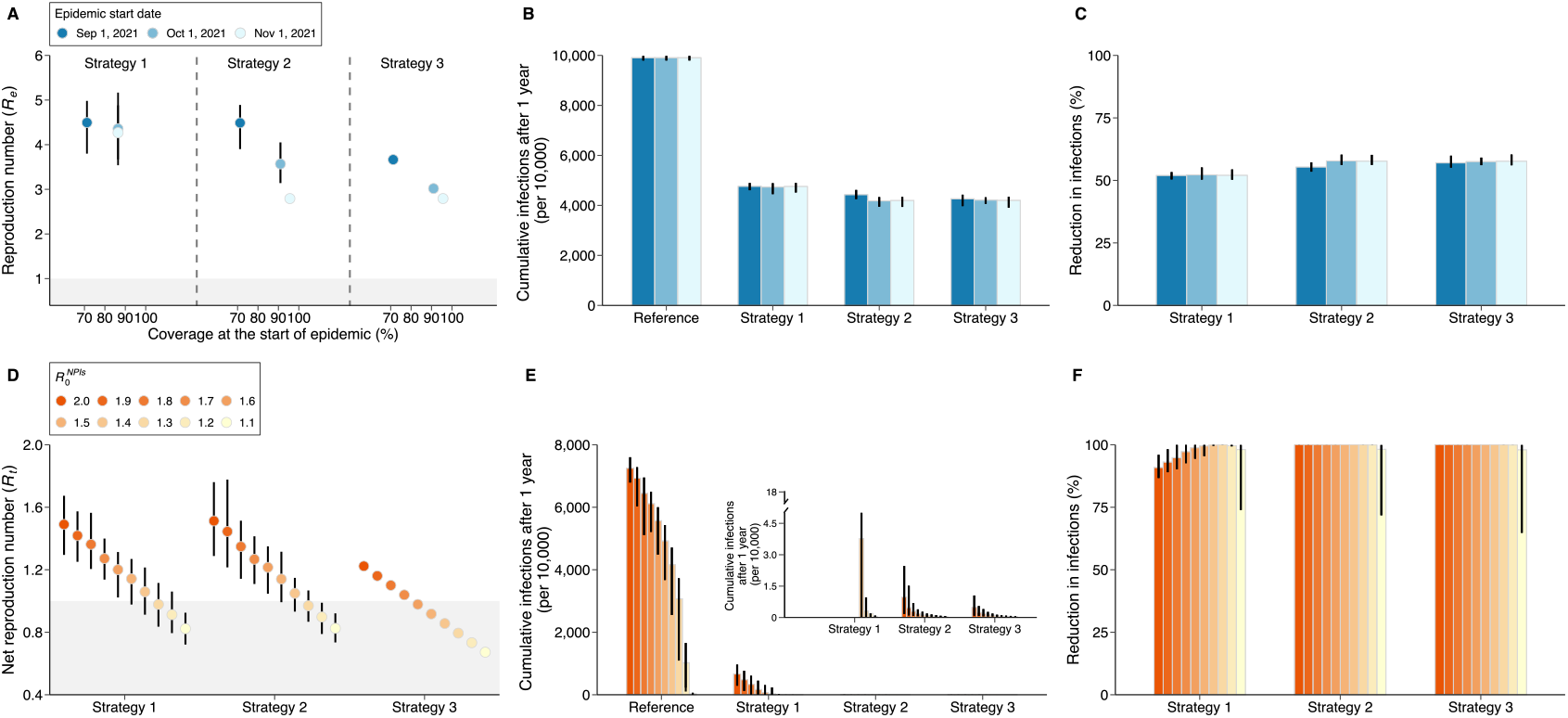
Impact of delaying the start of the epidemic and adopting NPIs. **A** Estimated effective reproduction number (*R*_*e*,_ mean and 95% CI) as a function of vaccine coverage at the time the infection is seeded (i.e., September 1, October 1 and November 1). Colors refer to the scenario of delaying the start of the epidemic to different date. The shaded area in gray indicates *R*_*e*_ ≤1. **B** Cumulative number of infections per 10,000 individuals after 1 simulated year for *reference scenario* and three vaccination strategies (mean and 95% CI). *Reference scenario* indicates no vaccination and no NPIs with *R*_*0*_=6.0 at the beginning of transmission. **C** Reduction in infections (mean and 95% CI) with respect to the *reference scenario*. **D** As A, but for estimated net reproduction number (*R*_*t*,_ mean and 95% CI) adopting different intensity of NPIs, 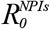. **E** As B, but for the scenario of adopting different intensity of NPIs. **F** As C, but for the scenario of adopting different intensity of NPIs.

### Scenario 2: Adopting NPIs in case of a new outbreak

The results presented so far suggest that herd immunity against Delta variant is not achievable at any time point. Adopting NPIs as a response to an epidemic outbreak can lower the transmission potential of the virus. It is thus worth investigating the synergetic effect of vaccination programs combined with NPIs of different intensity. It is important to note that we do not explicitly model every single measure to limit transmission (e.g., case isolation, contact tracing, wearing masks, social distancing, improved hygiene). These measures are implicit as concerted strategies that result in a decreased reproduction number. We define the value of the reproduction number in a fully susceptible population and under a certain level of NPIs as 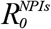. We explored 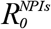 in the range 1.1-6.0. Values between 1 and 2 are showed in the main text, while larger values are shown in *SI Appendix* (Fig. S12).

The mean net reproduction number (which accounts both for immunity and interventions) on September 1, 2021 for strategy 1 and 2 can be reduced to below 1 only when 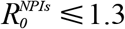, while for strategy 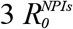 can be up to 1.6 (Fig. 3D). By forward vaccinating and simulating 1 year of epidemic, substantial infections could be reduced (close to 100% for 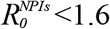) thanks to the synergetic effect of vaccination and NPIs (Fig. 3E-F and Fig. S13 in *SI Appendix*).

### Scenario 3: Delaying the start of the epidemic and adopting NPIs

To further improve the potential for vaccination-induced herd immunity and reduce COVID-19 burden, here we tested the combination of the two scenarios mentioned above: delaying the start of the epidemic and adopting NPIs of different level of intensity in response to a new outbreak.

Should an epidemic start in October-November 2021 and moderate NPIs (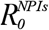 in the range 1.5-2.0) are adopted, strategy 2 and 3 can succeed in blocking transmission (Fig. 4B-C). In the case of strategy 1 (i.e., the vaccination policy excluding underage individuals), strict NPIs 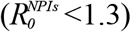 should be adopted to prevent a major epidemic wave (Fig. 4A and Fig. S14 in *SI Appendix*).

**Figure 4.**
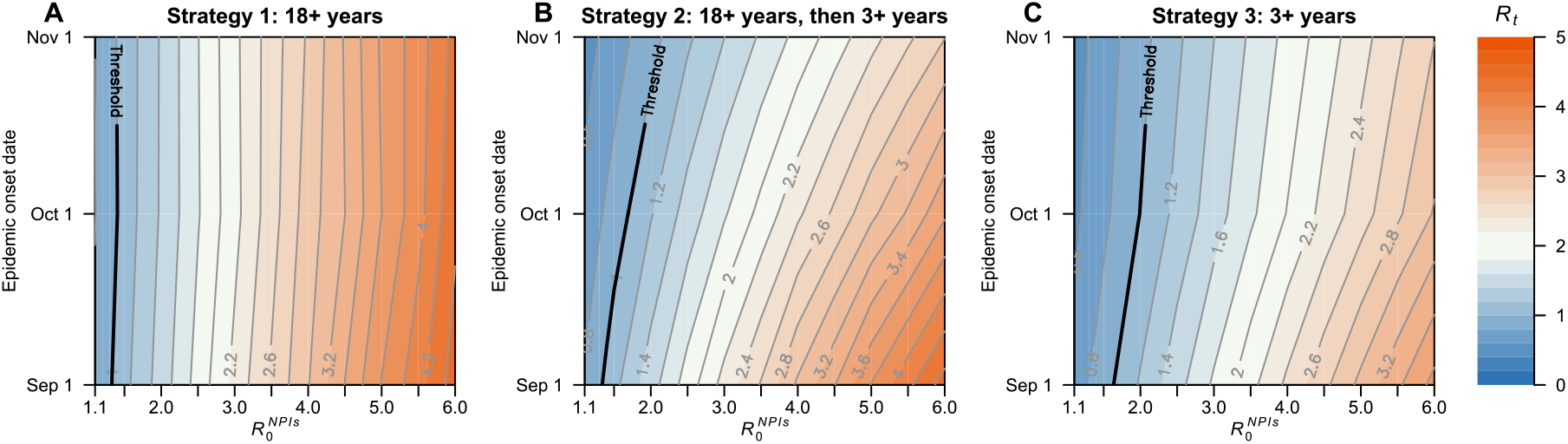
Impact of delaying the start of the epidemic start and adopting NPIs on estimated net reproduction number *R*_*t*_. **A** Estimated net reproduction number (*R*_*t*_) as a function of 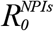 and epidemic start date for strategy 1. The bold line in black indicates the herd immunity threshold *R*_*t*_ =1. **B** As A, but for strategy 2. **C** As A, but for strategy 3.

The effectiveness of age-targeted vaccination strategies depend on the age-mixing patterns of the population ^12^. To test the robustness of our findings, we tested an alternative contact matrix for China and we found consistent results (Fig. S15-16 in *SI Appendix*).

### Herd immunity threshold

Till now, herd immunity is unattainable for any vaccination strategy considering the relatively low efficacy (54.3%) of the analyzed vaccine in preventing the infection from the Delta. We thus explored the potential of herd immunity for the three vaccination strategies given a higher efficacy (95%) (Fig. S17).

We estimated that *R*_*e*_ decreases below 1.0 only for strategy 2 and 3 (Fig. S17A). The estimated herd immunity threshold under these two strategies is 94.0% and 88.1% respectively, which suggests that level of immunity needed to lead the effective reproduction number below the epidemic threshold is lower if vaccination is extended to individuals aged 3-17 years early on.

We also estimated the infection attack rate under different vaccination coverages under the assumption that vaccination stops at the time the epidemic is seeded. This purely hypothetical scenario shows that when the adult population (18+ years of age) is vaccinated (strategy 1), despite a fairly high estimated reproduction number (2.9), the estimated infection attack rate is relatively low (13.2%) (Fig. S17B). In fact, given the age-targeted vaccination program and the lack of natural immunity, the susceptible population is mostly concentrated in the young population. The high number of contacts in younger age groups, combined with the high vaccination coverage in the rest of the population, lead to a fairly high reproduction number but, at the same time, the infections are focused on a small segment of the population only (young individuals) and thus the overall infection attack rate remains fairly low.

We also explored whether herd immunity is achievable or not and what is the herd immunity threshold by estimating *R*_*e*_ under the assumption of having access to a vaccine with a different efficacy (60%-100%) and exploring different scenarios on the vaccine coverage.

Our results show that, for a vaccine with an efficacy lower than 85%, herd immunity is unattainable, even in the extreme case where the vaccine coverage is 100% (Fig. 5). Vaccine-induced herd immunity may only be achievable with higher VE and high coverage. For example, for a vaccine with 90% efficacy against infection from the Delta variant, more than 93% of the population would need to be vaccinated to reach herd immunity (Fig. 5). In the presence of NPIs, the net reproduction number can be reduced below the unit for lower vaccine efficacy and coverage values (Fig. S18 in *SI Appendix*).

**Figure 5.**
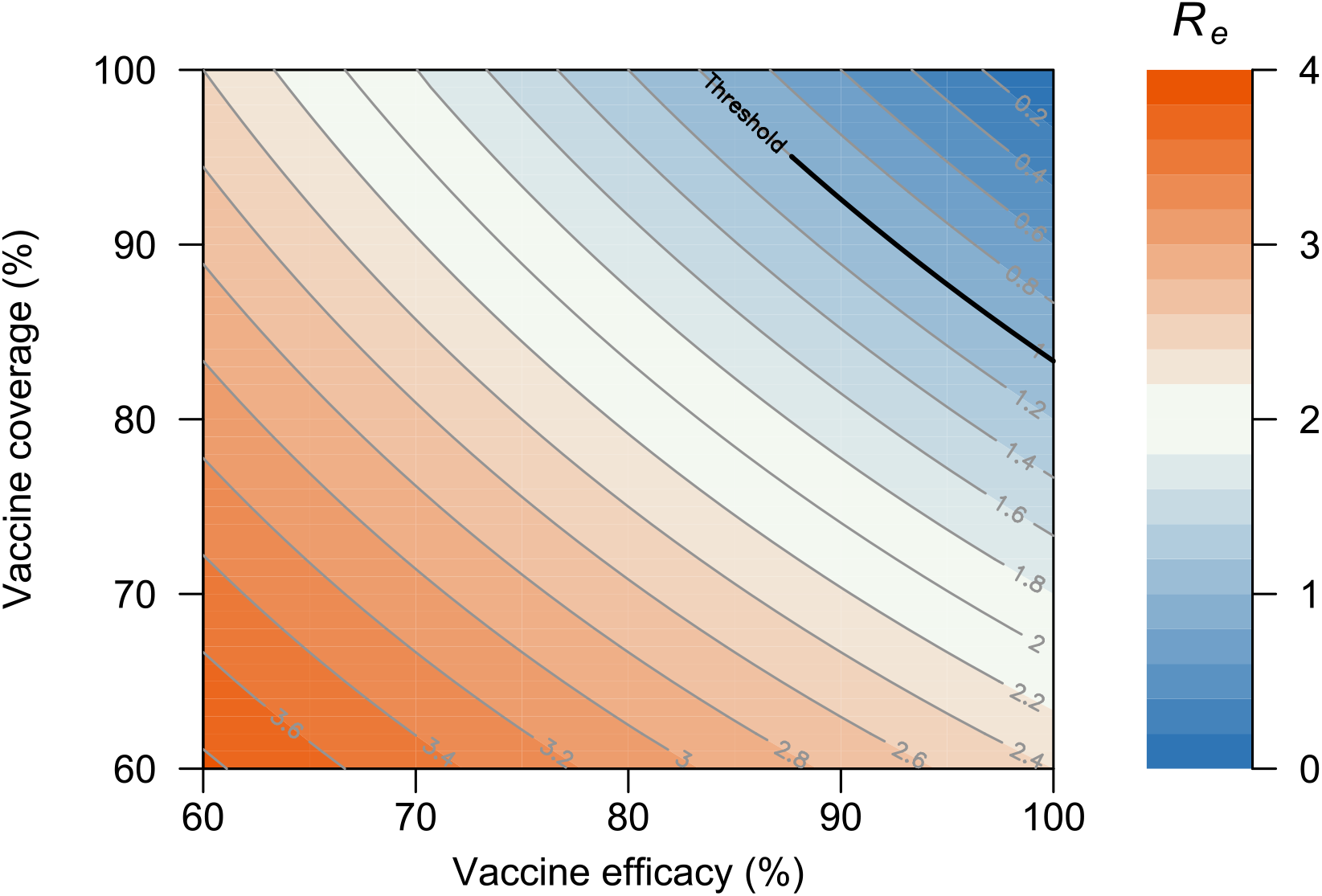
The impact of vaccine efficacy and vaccine coverage on estimated effective reproduction number *R*_*e*_. The bold line in black indicates the herd immunity threshold *R*_*e*_ =1. *R*_*e*_ is estimated in the scenario that all individuals are eligible to vaccination and vaccinated 2 doses before the epidemic starts.

## Discussion

Our study evaluates the feasibility of reaching herd immunity against the SARS-CoV-2 Delta variant through vaccination, considering heterogeneity in population age structure, age-specific contact patterns, vaccine efficacy, as well as biological characteristics of SARS-CoV-2. Our findings show that herd immunity is unlikely to be reached against the Delta variant given the relatively low efficacy of the current vaccines (developed against the historical SARS-CoV-2 lineage), also in the presence of prior natural immunity up to 30%. Even considering vaccines with higher efficacy, our results show the key role of extending the vaccination program to school-aged children in order to increase the potential of reaching herd immunity and reduce the infection attack rate. If we consider a protection against the Delta variant of 90% (which goes beyond current vaccines), herd immunity would require the vaccination of 93% of the whole population. The adoption of NPIs could prevent the spread of a major epidemic wave even when the herd immunity level is not reached, but such an option obviously entails social and economic costs. Further, all strategies considered in this study would mitigate the overwhelming majority of infections.

Our study explored if and when vaccination-induced herd immunity can be reached in China. Under the hypotheses that the circulating strain has the same transmissibility as Delta variant and that the vaccination campaign will not slow down due to vaccine hesitancy, herd immunity seems to remain unreachable even in the extreme case where the vaccine coverage is 100%. Nonetheless, it is important to remark that the effectiveness of the vaccination program is impacted both by the natural immunity accumulated in the population (which is close to 0 in China as of July 2021) and the age structure of the population. In fact, in populations with a higher natural immunity level and a lower proportion of children, herd immunity may be achievable. Our findings point to the importance of adopting NPIs and/or self-precautionary measures until herd immunity is reached or the burden of the epidemic becomes manageable. These measures can either help delay the seeding of the infection (e.g., strict border control measures) or, should an epidemic start to unfold, mitigate its burden (e.g., social distancing, contact tracing, testing, wearing masks, hygiene practices, limiting contacts). However, questions remain about which NPIs need to be implement, their intensity, and timing. Future studies are needed to address these questions.

A key role to determine the success of a vaccination campaign is played by the willingness-to-vaccine of the population. According to previous surveys on COVID-19 vaccine hesitancy, vaccine acceptance in China was estimated to vary between 60.4% and 91.3% for general population aged 18 years and above ^13-16^, and may be even lower for older adults ^16^ and healthcare workers ^17^. Similar estimates were obtained for several other countries including the UK (71.5%) ^18^ and the US (75.4%) ^18^. Given these levels of vaccine hesitancy, achieving high levels of coverage may remain an elusive target. Efforts to increase population’s confidence and willingness to be vaccinated will thus be of paramount importance to allow a return to a pre-COVID-19 lifestyle. Our study shows that the spread of the more transmissible Delta variant has substantially increased the herd immunity threshold to a level that may not be feasible in any population, so that mitigation strategies become even more relevant.

Previous studies have estimated the herd immunity threshold either through natural infection or vaccination under the assumption of an homogenously mixed population ^19-21^, but heterogeneity in contact structure, age structure of the population, susceptibility to infection by age, and order in which individuals are vaccinated are all key factors shaping the herd immunity level ^8^. The developed model is based on social mixing patterns estimated for the Shanghai population ^9^ and on China-specific data on COVID-19 epidemiology (population immunity, etc.). Nevertheless, the introduced modeling framework is flexible and can be tailored to other countries. We tested a scenario somehow resembling the situation in the USA, where we considered naturally immunity and the adoption of BNT162b2/Pfizer vaccine, whose efficacy against the Delta variant was estimated at 79%. Also, in this scenario, we estimate that herd immunity may not be reached (Fig. S19 in *SI Appendix*). Moreover, vaccination hesitancy may jeopardize the vaccination effort in the US and other Western countries as well.

This study is prone to the limitations pertaining to modeling exercises. First, VE against infections from the Delta variant was inferred instead of directly measures from epidemiological observations. Moreover, VE for children have not been estimated for the vaccines in use in China; therefore, we have assumed the same VE as in adults based on immunogenicity studies ^22^. Given such a lack of field evidence, we have conducted a sensitivity analysis where a lower vaccine efficacy is assumed for children. The overall conclusions of the study do not change. Still, further data on age-specific vaccine efficacy could help refine the obtained estimates.

Second, we assumed that immunity induced either from infection or vaccination lasts more than the time horizon considered in the simulations (i.e., 1 year). There are both evidence from laboratory studies and the field suggesting that the protection lasts several months ^23^. Despite these preliminary pieces of evidence, the duration of the immunity remains a research area of paramount importance and intrinsically linked to viral evolution. It is also possible that waning immunity will continue to provide protection against severe disease but only partial against infection or transmission, which affects the herd immunity threshold. Overall, the duration and quality of immunity will determine the periodicity of COVID-19 outbreaks globally ^24,25^. Third, in the baseline scenario, we referred to an inactivated SARS-CoV-2 vaccine (BBIBP-CorV) taken to be 54.3% efficacious against the Delta variant infection. However, several other vaccines (including CoronaVac, WBIP-CorV, Ad5-nCoV, and ZF2001) are licensed and have been used in China. We varied vaccine efficacy up to 79% in sensitivity analyses. The main conclusion about the potential of herd immunity and the need to extend the vaccination campaign to children as well as to use more efficacious vaccines is unaltered.

In conclusion, based on the current evidence, reaching vaccine-induced herd immunity in a population with little/no natural immunity is challenging. Key steps will be the authorization of COVID-19 vaccines for children, and minimize vaccine hesitancy. These, together with highly efficacious vaccines or booster vaccinations, will be even more crucial given the possible emergence of new, even more transmissible, SARS-CoV-2 variants. Importantly, even if herd immunity is unlikely to be reached, vaccination will still dramatically reduce COVID-19 burden.

## Data Availability

The availability of all data can be addressed to the correspondence author.

## Contributors

H.Y. conceived and designed the study. H.Y. and M.A. supervised the study. M.A. designed the model. H.L. developed the model. H.L., J.Z., J.C., J.Y., X.D. analyzed the model outputs. H.L., C.P., X.D, and Z.C. prepared the tables and figures. H.L. and J.Z. prepared the first draft of the manuscript. W.Z., Q.W. and X. C. participated in data collection. X.C and Z.C. updated the relative literatures. H.Y., M.A. W.Z. and C. V. revised the content critically. All authors contributed to review and revision and approved the final manuscript as submitted and agree to be accountable for all aspects of the work.

## Disclaimer

This article does not necessarily represent the views of the NIH or the US government.

## Acknowledgments

The study was supported by grants from the Key Program of the National Natural Science Foundation of China (82130093), European Union Grant 874850 MOOD (MOOD 000), and the National Institute for Health Research (NIHR) (grant no. 16/137/109) using UK aid from the UK Government to support global health research. We also acknowledge grant from Shanghai Key Laboratory of Infectious Diseases and Biosafety Emergency Response (20dz2260100). The views expressed in this publication are those of the author(s) and not necessarily those of the NIHR or the UK Department of Health and Social Care.

## Data sharing

The data and code that support the findings of this study will be made available in GitHub upon manuscript acceptance.

