## supplemental files for "Herd immunity induced by COVID-19 vaccination programs and suppression of epidemics caused by the SARS-CoV-2 Delta variant in China"

#### 1 SARS-CoV-2 transmission and vaccination model

##### *Transmission and vaccination model*

We developed an age-stratified stochastic susceptible-infectious-removed model of SARS-CoV-2 transmission. The population is divided into 17 age groups with 16 5-year age groups from 0 to 79 years and 1 age group for individuals aged 80 years or older. Age-mixing patterns were derived from a contact survey study conducted in Shanghai in 2017-2018 [1]. The model is used to simulate a set of vaccination programs.

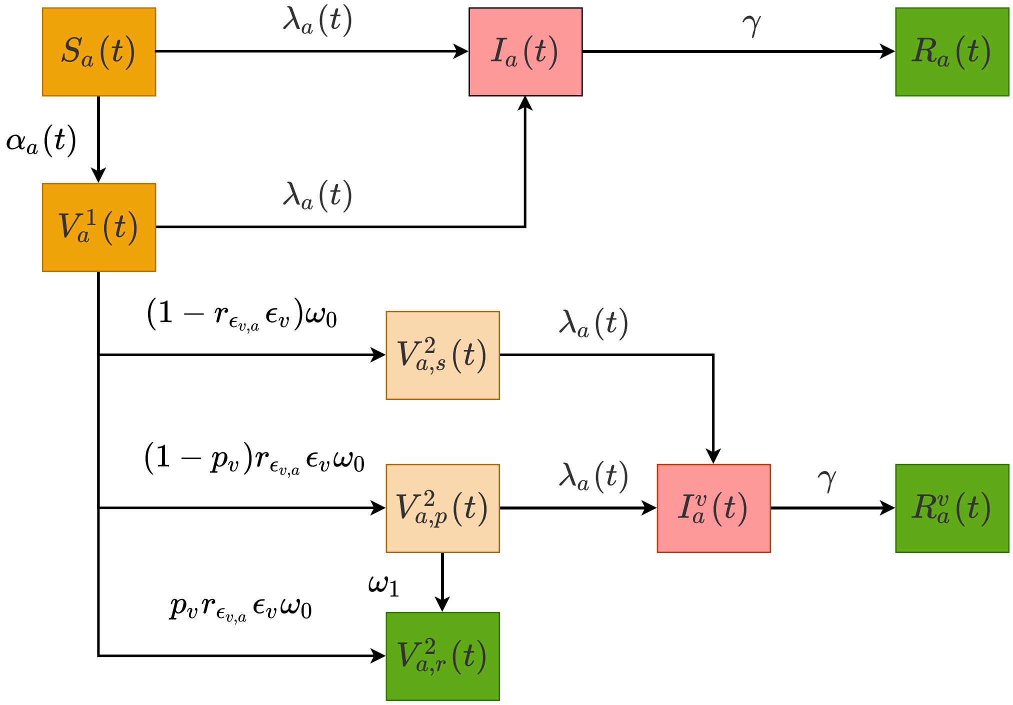

**Figure S1. Schematic figure of SARS-CoV-2 transmission and vaccination model.** Compartments used in the model include (take age group $a \in [1, 17]$ at time $t$ as an example): unvaccinated susceptible individuals $S_a(t)$, unvaccinated infected individuals $I_a(t)$, unvaccinated removed individuals $R_a(t)$, individuals who have received one vaccine dose $V_a^1(t)$, unprotected vaccinated individuals who have received two doses $V_{a,s}^2(t)$, temporarily unprotected vaccinated individuals who have received two doses and will gain protection 14 days later $V_{a,p}^2(t)$, protected vaccinated individuals who have received two doses $V_{a,r}^2(t)$, vaccinated infected individuals $I_a^V(t)$, and vaccinated removed individuals $R_a^V(t)$. Note that vaccinated individuals are defined as those administered 2 doses. Parameters used in the model include: time- and age-dependent force of infection $\lambda_a(t)$, recovery rate from infection $\gamma$, relative vaccine efficacy against infection right after administration of 2nd dose $p_v$ as compared with maximum protection (i.e., after the ramp-up of the 2nd dose) $\epsilon_v$, and vaccine efficacy of age group $a$ relative to adults aged 18-59 years ($r_{\epsilon_v,a}$), probability of administration of 1st dose ($\alpha_a$), time interval between administration of first and second dose ($1/\omega_0$) and maximum protection ($1/\omega_1$).

The SARS-CoV-2 transmission and vaccination model is schematically represented in Figure S1 and it is described by the following differential equations:

$$\dot{S_a} = -\lambda_a S_a - \alpha_a S_a \tag{1}$$

$$\dot{I_a} = \lambda_a S_a + \lambda_a V_a^1 - \gamma I_a \tag{2}$$

$$\dot{R_a} = \gamma I_a \tag{3}$$

$$\dot{V_a^1} = \alpha_a S_a - \lambda_a V_a^1 - \omega_0 V_a^1 \tag{4}$$

$$\dot{V_{a,s}^2} = \left(1 - r_{\epsilon_{v,a}} \epsilon_v\right)\omega_0 V_a^1 - \lambda_a V_{a,s}^2 \tag{5}$$

$$\dot{V_{a,p}^2} = (1 - p_v) r_{\epsilon_{v,a}} \epsilon_v \omega_0 V_a^1 - \lambda_a V_{a,p}^2 - \omega_1 V_{a,p}^2 \tag{6}$$

$$\dot{V_{a,r}^2} = p_v r_{\epsilon_{v,a}} \epsilon_v \omega_0 V_a^1 + \omega_1 V_{a,p}^2 \tag{7}$$

$$\dot{I_a^V} = \lambda_a V_{a,s}^2 + \lambda_a V_{a,p}^2 - \gamma I_a^V \tag{8}$$

$$\dot{R_a^V} = \gamma I_a^V \tag{9}$$

where:

- $S_a$ represents the unvaccinated susceptible individuals.
- $I_a$ represents the unvaccinated infected individuals.
- $R_a$ represents the unvaccinated removed individuals.
- $V_a^1$ represents the individuals who have received one vaccine dose.

- $V_{a,s}^2$ represents the unprotected vaccinated individuals who have received two doses.
- $V_{a,p}^2$ represents the temporarily unprotected vaccinated individuals who have received two doses and will gain protection 14 days later.
- $V_{a,r}^2$ represents the protected vaccinated individuals who have received two doses.
- $I_a^V$ represents the vaccinated infected individuals.
- $R_a^V$ represents the vaccinated removed individuals.
- $\lambda_a$ represents the time- and age-dependent force of infection.
- $\gamma$ represents the recovery rate, which corresponds to the length of the generation time in an SIR model [2]; we consider an average generation time of 7 days [3].
- $\epsilon_v$ represents the maximum protection (i.e., after the ramp-up of the 2nd dose).
- $p_v$ represents relative vaccine efficacy against infection right after administration of 2nd dose as compared with maximum protection.
- $r_{\epsilon_{v,a}}$ represents vaccine efficacy of age group $a$ relative to adults aged 18-59 years.
- $\alpha_a$ represents the probability of administration of 1st dose.
- $1/\omega_0$ represents time interval between administration of first and second dose.
- $1/\omega_1$ represents maximum protection.

The time- and age-dependent force of infection $\lambda_a(t)$ to which susceptible individuals of age group $a$ are exposed to SARS-CoV-2 infection is defined as:

$$\lambda_a(t) = \beta\varepsilon_a \sum_{\tilde{a}} C_{a,\tilde{a}} \frac{I_{\tilde{a}}+I_{\tilde{a}}^V}{N_{\tilde{a}}} \tag{10}$$

Where:

- $\beta$ is a scaling factor shaping SARS-CoV-2 transmissibility.

- $\varepsilon_a$ is the relative susceptibility to SARS-CoV-2 infection of age group $a$.
- $C_{a,\tilde{a}}$ represents the age-group-specific contact matrix, whose elements describe the daily mean numbers of that a person in age group $a$ has with individuals in age group $\tilde{a}$.
- $N_a$ represents the number of individuals of age group $a$.

Considering the daily vaccination capacity (see Sec. 5 of *SI Appendix*), we prioritize those individuals who need to receive the 2$^{nd}$ dose at time $t$, then the remaining daily capacity is randomly allocated to those who are eligible to receive the 1$^{st}$ dose. At each time $t$, the first dose of vaccinations is administrated to a fraction of susceptible individuals in the population in age group $a$:

$$\alpha_a = \frac{d_a(t)}{S_a(t)} \tag{11}$$

where $d_a(t)$ represents the number of first doses to be allocated to individuals in age group $a$ at time $t$ under the designed vaccination strategies.

A summary of model parameters is reported in Table S1.

**Table S1.** Summary of parameters used to model Delta strain.

| **Description** | **Baseline analysis** | **Sensitivity analysis** |
|---|---|---|
| **Epidemiology** | | |
| Average generation time $(\mathbf{1/\gamma})$ | 7 days (95% CI: 3.6-11.3) [3] | - |
| Basic reproduction number $(\boldsymbol{R_0})$ | 6.0 [a 4-17] | - |

| | | |
|---|---|---|
| reproduction number in a fully susceptible population and under certain NPIs ($\boldsymbol{R_0^{NPIs}}$) | 1.1-6.0 | - |
| Initially infected individuals | 40 [18] | 10, 20, 100 |
| Initial natural immunity | 0 [19] | 10%, 20%, 30% |
| Relative susceptibility to infection by age ($\boldsymbol{\varepsilon_a}$) | 0.58 (95% CI 0.34-0.98) for $a < 15$;<br><br>1 for $15 \le a < 65$;<br><br>1.65 (95% CI 1.03-2.65) for $a \ge 65$ [20]. | Homogeneous susceptibility |
| Epidemic start date | Sep 1, 2021 | Oct 1, 2021, Nov 1, 2021 |
| **Vaccination** | | |
| Interval between the administration of 1st and 2nd dose ($\boldsymbol{1/\omega_0}$) | 21 days [21] | 14 days |
| Delay between the administration of the 2nd dose of vaccination and maximum protection ($\boldsymbol{1/\omega_1}$) | 14 days [21] | - |
| Vaccine efficacy in preventing the infection for individuals aged 18-59 years ($\boldsymbol{\epsilon_v}$) | 54.3% [b 21-23] | 0.6, 0.79 |
| Relative vaccine efficacy against infection right after administration of 2nd dose as compared with maximum protection ($\boldsymbol{p_v}$) | 83.8% [c 24] | - |
| Relative vaccine efficacy for individuals aged < 18 and > 59 years as compared to individuals aged 18-59 years ($\boldsymbol{r_{\epsilon_v,a}}$) | 100% | 75%, 50% |

| | | |
|---|---|---|
| Daily vaccination capacity | See section 5 | - |

[a] Estimated based on the conclusion [4-7] that the $R_0$ of wild strain is 2.5, and transmissibility of Alpha variant is 60% [8-14] higher as compared with that of wild strain, and transmissibility of Delta variant is 50% [15-17] higher as compared with that of Alpha variant.

[b] Following the work by *Khoury et al* [23], we estimate vaccine protection against Delta variant infections by using two factors: i) the estimated efficacy against the infection from the historical lineages for BBIBP-CorV vaccine [21], and ii) the reduction of neutralizing antibodies for the Delta variant estimated from *in vitro* neutralization assay [22]. Specifically, first, using the equations $E_I\ (n|n_{50}, k) = \frac{1}{1+e^{-k(n-n_{50})}}$, we modelled the relationship between efficacy against the original lineage and neutralizing titers (see Khoury et al. [23] for details). Then, the distributions of changes in neutralizing antibodies for the Delta variants were added into the equation $P\ (n_{50}, k, \mu_s, \sigma_s) = \int_{-\infty}^{+\infty} \mathrm{E}_I\ (n|n_{50}, k)\ f(n|\mu_s, \sigma_s) dn$ to estimate vaccine efficacy (see Khoury et al. [23] for details).

[c] We assumed that individuals could be protected with a lower efficacy within 0-13 days after the second dose and the ratio is the same for both historical lineages and Delta strain. In order to estimate the efficacy over time for BBIBP-Corv, we refer to another inactivated vaccine used in China (i.e., CoronaVac with 2 doses and 14 days apart). The efficacy for 0-13 days after second-dose and 14 days after second-dose are 42.5% and 50.7%, respectively [24]. Thus, the efficacy within 0-13 days after second-dose in our main analysis is inferred as 54.3% * (42.5%/50.7%) = 45.5%, while we assume that there is no protection before 14 days since the administration of the second dose for sensitivity analysis.

#### 2 Transmission rate and basic reproduction number in the absence of NPIs

The basic reproduction number $R_0$ with age structure can be computed by using the Next-Generation matrix (NGM) approach [25] as:

$$R_0 = \frac{\beta}{\gamma}\rho(K_1) \tag{12}$$

Where $\rho(K_1)$ presents the dominant eigenvalue of combined matrix $K_1$,

$$K_1 = \begin{bmatrix} \varepsilon_1 C_{11}\frac{N_1}{N_1} & \cdots & \varepsilon_1 C_{1n}\frac{N_1}{N_n} \\ \vdots & \ddots & \vdots \\ \varepsilon_n C_{n1}\frac{N_n}{N_1} & \cdots & \varepsilon_n C_{nn}\frac{N_n}{N_n} \end{bmatrix}_{17} \tag{13}$$

Thus, infectious rate $\beta$ can be computed based on equation from (12) as

$$\beta = \frac{\gamma \times R_0}{\rho(K_1)} \tag{14}$$

#### 3 Estimation of the effective reproduction number ($R_e$)

The effective reproduction number $R_e$ accounts for the immunity in the population (either natural or from vaccination). It can be computed by using the Next-Generation matrix (NGM) approach [25] as follows:

$$R_v = \frac{\beta}{\gamma}\rho(K_2) \tag{15}$$

Where $\rho(K_2)$ presents the dominant eigenvalue of combined matrix $K_2$,

$$K_2 = \begin{bmatrix} \varepsilon_1 C_{11}\frac{N_1 - N_1 * c_1 * \epsilon_v}{N_1} & \cdots & \varepsilon_1 C_{1n}\frac{N_1 - N_1 * c_1 * \epsilon_v}{N_n} \\ \vdots & \ddots & \vdots \\ \varepsilon_n C_{n1}\frac{N_n - N_n * c_n * \epsilon_v}{N_1} & \cdots & \varepsilon_n C_{nn}\frac{N_n - N_n * c_n * \epsilon_v}{N_n} \end{bmatrix}_{17} \tag{16}$$

Where $c_a$ represents the vaccine coverage of age group $a$ and $\epsilon_v$ represents the vaccine efficacy in preventing the infection.

#### 4 Vaccine contraindication and pregnant women

All individuals with vaccine contraindications and pregnant women are excluded from the target population of the simulated vaccination programs. Vaccine contraindications are defined according to the guideline reported by WHO [26-29]. The proportion of individuals with vaccine contraindications and pregnant women in Chinese population is listed in Table S2. We generated the proportion of individuals with at least 1 underlying condition of age group $a$, $P_a$, by using the following equation

$$P_a = 1 - [(1 - p(\mathrm{C}_1))(1 - p(\mathrm{C}_2)) \times \ldots \times (1 - p(\mathrm{C}_n)] \quad (17)$$

Where $C_n$ $(n = 1, 2, 3, \ldots, 37)$ represents the underly conditions listed in Table S2 and $p(C_n)$ represents the proportion of individuals with underlying condition $C_n$. We then used the ratio between real and estimated probability of individuals with at least 1 underlying condition reported in the literatures (68.2%) [30-32] to adjust probability of each group.

**Table S2.** The proportion of pregnant women and vaccine contraindications by age groups.

| Proportion (%) | Age group (years) | | | | | | | | | | | | | | | | | |
|---|---|---|---|---|---|---|---|---|---|---|---|---|---|---|---|---|---|---|
| | 0-4 | 5-9 | 10-14 | 15-19 | 20-24 | 25-29 | 30-34 | 35-39 | 40-44 | 45-49 | 50-54 | 55-59 | 60-64 | 65-69 | 70-74 | 75-79 | 80-84 | 85+ |
| Pregnant women | 0.0 | 0.0 | 0.0 | 0.7 | 5.3 | 10.3 | 5.2 | 2.6 | 0.7 | 0.3 | 0.0 | 0.0 | 0.0 | 0.0 | 0.0 | 0.0 | 0.0 | 0.0 |
| Contraindications | 0.2 | 0.1 | 0.1 | 0.1 | 0.2 | 0.2 | 0.3 | 0.5 | 0.7 | 0.9 | 1.2 | 1.7 | 2.2 | 2.7 | 3.2 | 3.2 | 2.8 | 2.2 |
| Liver cancer due to NASH | 0.0 | 0.0 | 0.0 | 0.0 | 0.0 | 0.0 | 0.0 | 0.0 | 0.0 | 0.0 | 0.0 | 0.0 | 0.0 | 0.0 | 0.0 | 0.0 | 0.0 | 0.0 |
| Colon and rectum cancer | 0.0 | 0.0 | 0.0 | 0.0 | 0.0 | 0.0 | 0.1 | 0.1 | 0.1 | 0.2 | 0.3 | 0.4 | 0.6 | 0.8 | 1.0 | 1.0 | 0.9 | 0.7 |
| Lip and oral cavity cancer | 0.0 | 0.0 | 0.0 | 0.0 | 0.0 | 0.0 | 0.0 | 0.0 | 0.0 | 0.0 | 0.0 | 0.0 | 0.0 | 0.0 | 0.0 | 0.0 | 0.0 | 0.0 |
| Nasopharynx cancer | 0.0 | 0.0 | 0.0 | 0.0 | 0.0 | 0.0 | 0.0 | 0.1 | 0.1 | 0.1 | 0.1 | 0.1 | 0.1 | 0.1 | 0.1 | 0.0 | 0.0 | 0.1 |
| Other pharynx cancer | 0.0 | 0.0 | 0.0 | 0.0 | 0.0 | 0.0 | 0.0 | 0.0 | 0.0 | 0.0 | 0.0 | 0.0 | 0.0 | 0.0 | 0.0 | 0.0 | 0.0 | 0.0 |
| Gallbladder and biliary tract | 0.0 | 0.0 | 0.0 | 0.0 | 0.0 | 0.0 | 0.0 | 0.0 | 0.0 | 0.0 | 0.0 | 0.0 | 0.0 | 0.0 | 0.0 | 0.0 | 0.0 | 0.0 |
| Pancreatic cancer | 0.0 | 0.0 | 0.0 | 0.0 | 0.0 | 0.0 | 0.0 | 0.0 | 0.0 | 0.0 | 0.0 | 0.0 | 0.0 | 0.0 | 0.0 | 0.0 | 0.0 | 0.0 |
| Malignant skin melanoma | 0.0 | 0.0 | 0.0 | 0.0 | 0.0 | 0.0 | 0.0 | 0.0 | 0.0 | 0.0 | 0.0 | 0.0 | 0.0 | 0.0 | 0.0 | 0.0 | 0.0 | 0.0 |
| Ovarian cancer | 0.0 | 0.0 | 0.0 | 0.0 | 0.0 | 0.0 | 0.0 | 0.0 | 0.0 | 0.0 | 0.0 | 0.0 | 0.0 | 0.0 | 0.0 | 0.0 | 0.0 | 0.0 |
| Testicular cancer | 0.0 | 0.0 | 0.0 | 0.0 | 0.0 | 0.0 | 0.0 | 0.0 | 0.0 | 0.0 | 0.0 | 0.0 | 0.0 | 0.0 | 0.0 | 0.0 | 0.0 | 0.0 |
| Kidney cancer | 0.0 | 0.0 | 0.0 | 0.0 | 0.0 | 0.0 | 0.0 | 0.0 | 0.0 | 0.0 | 0.0 | 0.0 | 0.0 | 0.0 | 0.1 | 0.0 | 0.0 | 0.0 |
| Bladder cancer | 0.0 | 0.0 | 0.0 | 0.0 | 0.0 | 0.0 | 0.0 | 0.0 | 0.0 | 0.0 | 0.0 | 0.1 | 0.1 | 0.1 | 0.2 | 0.2 | 0.1 | 0.1 |
| Brain and central nervous | 0.0 | 0.0 | 0.0 | 0.0 | 0.0 | 0.0 | 0.0 | 0.0 | 0.0 | 0.0 | 0.0 | 0.0 | 0.0 | 0.0 | 0.0 | 0.0 | 0.0 | 0.0 |
| Thyroid cancer | 0.0 | 0.0 | 0.0 | 0.0 | 0.0 | 0.0 | 0.0 | 0.0 | 0.0 | 0.0 | 0.0 | 0.0 | 0.0 | 0.0 | 0.0 | 0.0 | 0.0 | 0.0 |
| Mesothelioma | 0.0 | 0.0 | 0.0 | 0.0 | 0.0 | 0.0 | 0.0 | 0.0 | 0.0 | 0.0 | 0.0 | 0.0 | 0.0 | 0.0 | 0.0 | 0.0 | 0.0 | 0.0 |
| Larynx cancer | 0.0 | 0.0 | 0.0 | 0.0 | 0.0 | 0.0 | 0.0 | 0.0 | 0.0 | 0.0 | 0.0 | 0.0 | 0.1 | 0.1 | 0.1 | 0.1 | 0.1 | 0.0 |
| Tracheal, bronchus, and lung | 0.0 | 0.0 | 0.0 | 0.0 | 0.0 | 0.0 | 0.0 | 0.0 | 0.0 | 0.0 | 0.1 | 0.1 | 0.2 | 0.3 | 0.4 | 0.4 | 0.4 | 0.3 |
| Breast cancer | 0.0 | 0.0 | 0.0 | 0.0 | 0.0 | 0.0 | 0.1 | 0.2 | 0.3 | 0.3 | 0.4 | 0.5 | 0.6 | 0.6 | 0.6 | 0.6 | 0.5 | 0.5 |

| | | | | | | | | | | | | | | | | | | |
|---|---|---|---|---|---|---|---|---|---|---|---|---|---|---|---|---|---|---|
| Cervical cancer | 0.0 | 0.0 | 0.0 | 0.0 | 0.0 | 0.0 | 0.0 | 0.1 | 0.1 | 0.1 | 0.1 | 0.1 | 0.1 | 0.0 | 0.0 | 0.0 | 0.0 | 0.0 |
| Uterine cancer | 0.0 | 0.0 | 0.0 | 0.0 | 0.0 | 0.0 | 0.0 | 0.0 | 0.0 | 0.1 | 0.1 | 0.1 | 0.1 | 0.1 | 0.1 | 0.0 | 0.0 | 0.0 |
| Prostate cancer | 0.0 | 0.0 | 0.0 | 0.0 | 0.0 | 0.0 | 0.0 | 0.0 | 0.0 | 0.0 | 0.0 | 0.1 | 0.2 | 0.4 | 0.5 | 0.6 | 0.6 | 0.3 |
| Esophageal cancer | 0.0 | 0.0 | 0.0 | 0.0 | 0.0 | 0.0 | 0.0 | 0.0 | 0.0 | 0.0 | 0.0 | 0.1 | 0.1 | 0.1 | 0.2 | 0.2 | 0.2 | 0.1 |
| Stomach cancer | 0.0 | 0.0 | 0.0 | 0.0 | 0.0 | 0.0 | 0.0 | 0.0 | 0.1 | 0.1 | 0.1 | 0.2 | 0.3 | 0.3 | 0.4 | 0.4 | 0.4 | 0.2 |
| Liver cancer due to hepatitis B | 0.0 | 0.0 | 0.0 | 0.0 | 0.0 | 0.0 | 0.0 | 0.0 | 0.0 | 0.0 | 0.0 | 0.0 | 0.0 | 0.0 | 0.0 | 0.0 | 0.0 | 0.0 |
| Liver cancer due to hepatitis C | 0.0 | 0.0 | 0.0 | 0.0 | 0.0 | 0.0 | 0.0 | 0.0 | 0.0 | 0.0 | 0.0 | 0.0 | 0.0 | 0.0 | 0.0 | 0.0 | 0.0 | 0.0 |
| Liver cancer due to alcohol use | 0.0 | 0.0 | 0.0 | 0.0 | 0.0 | 0.0 | 0.0 | 0.0 | 0.0 | 0.0 | 0.0 | 0.0 | 0.0 | 0.0 | 0.0 | 0.0 | 0.0 | 0.0 |
| Liver cancer due to other | 0.0 | 0.0 | 0.0 | 0.0 | 0.0 | 0.0 | 0.0 | 0.0 | 0.0 | 0.0 | 0.0 | 0.0 | 0.0 | 0.0 | 0.0 | 0.0 | 0.0 | 0.0 |
| Myelodysplastic, | 0.0 | 0.0 | 0.0 | 0.0 | 0.0 | 0.0 | 0.0 | 0.0 | 0.0 | 0.1 | 0.1 | 0.2 | 0.3 | 0.4 | 0.4 | 0.4 | 0.4 | 0.3 |
| Acute lymphoid leukemia | 0.1 | 0.0 | 0.0 | 0.0 | 0.0 | 0.0 | 0.0 | 0.0 | 0.0 | 0.0 | 0.0 | 0.0 | 0.0 | 0.0 | 0.1 | 0.0 | 0.0 | 0.0 |
| Chronic lymphoid leukemia | 0.0 | 0.0 | 0.0 | 0.0 | 0.0 | 0.0 | 0.0 | 0.0 | 0.0 | 0.0 | 0.0 | 0.0 | 0.0 | 0.0 | 0.0 | 0.0 | 0.0 | 0.0 |
| Acute myeloid leukemia | 0.0 | 0.0 | 0.0 | 0.0 | 0.0 | 0.0 | 0.0 | 0.0 | 0.0 | 0.0 | 0.0 | 0.0 | 0.0 | 0.0 | 0.0 | 0.0 | 0.0 | 0.0 |
| Chronic myeloid leukemia | 0.0 | 0.0 | 0.0 | 0.0 | 0.0 | 0.0 | 0.0 | 0.0 | 0.0 | 0.0 | 0.0 | 0.0 | 0.0 | 0.0 | 0.0 | 0.0 | 0.0 | 0.0 |
| Other leukemia | 0.1 | 0.1 | 0.0 | 0.0 | 0.0 | 0.0 | 0.0 | 0.0 | 0.0 | 0.0 | 0.0 | 0.0 | 0.0 | 0.0 | 0.0 | 0.0 | 0.0 | 0.0 |
| Hodgkin lymphoma | 0.0 | 0.0 | 0.0 | 0.0 | 0.0 | 0.0 | 0.0 | 0.0 | 0.0 | 0.0 | 0.0 | 0.0 | 0.0 | 0.0 | 0.0 | 0.0 | 0.0 | 0.0 |
| Non-Hodgkin lymphoma | 0.0 | 0.0 | 0.0 | 0.0 | 0.0 | 0.0 | 0.0 | 0.0 | 0.0 | 0.0 | 0.0 | 0.0 | 0.1 | 0.1 | 0.1 | 0.1 | 0.1 | 0.1 |
| Multiple myeloma | 0.0 | 0.0 | 0.0 | 0.0 | 0.0 | 0.0 | 0.0 | 0.0 | 0.0 | 0.0 | 0.0 | 0.0 | 0.0 | 0.0 | 0.0 | 0.0 | 0.0 | 0.0 |
| Other malignant neoplasms | 0.1 | 0.0 | 0.1 | 0.1 | 0.0 | 0.0 | 0.0 | 0.0 | 0.1 | 0.1 | 0.1 | 0.1 | 0.2 | 0.2 | 0.3 | 0.3 | 0.2 | 0.2 |

#### 5 Vaccine administration capacity

We systematically collected information about COVID-19 vaccine administer in China from the press conferences of the Joint Prevention and Control Mechanism of the State Council [33]. The first data was recorded on November 30, 2020. At that time, the vaccination program was targeting all individuals aged between 18 and 59 years. On April 1, 2021, the target population was extended to include all individuals aged 18+ years. Table S3 shows the cumulative number of doses administered over time.

Since the number of administrated doses was not released every day, we interpolate the cumulative number of doses using an exponential model. The model is specified as,

$$N_t = N_0 e^{rt} \tag{18}$$

where, $N_0$ and $N_t$ represent the cumulative number of doses at time 0 and $t$, respectively, and $r$ is the growth rate.

The obtained fit until May 23, 2021 has a root mean square error (RMSE) of 8.298 (Figure S2 A). The increasing trend in the number of doses cannot continue to grow indefinitely. As such, we cupped the vaccination capacity at 17.53 million doses per day, as observed on June 2, 2021 (Fig. S2B). This capacity threshold (i.e., 17.53 million per day) corresponds to 294,234 doses per day for Shanghai (population of about 24 million) and is kept constant until the end of the simulation.

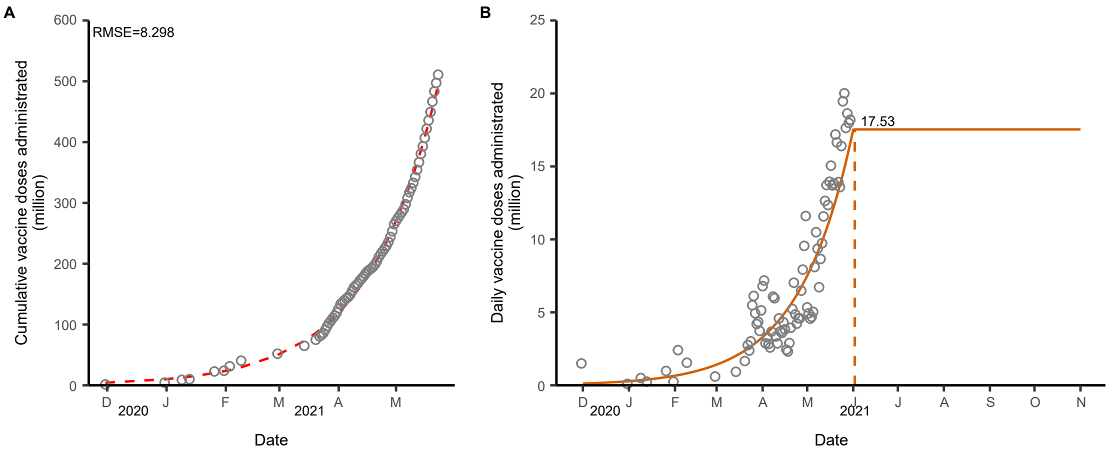

**Figure S2. Modelled vaccine capacity. A** Original and fitted cumulative number of doses administrated. **B** Projected daily vaccine capacity for the baseline scenario. Dots and lines represent empirical and fitted data.

**Table S3.** Cumulative number of COVID-19 doses administered in China.

| Date | Cumulative doses | Date | Cumulative doses |
|---|---|---|---|
| 2020-11-30 | 1,500,000 | 2021-04-17 | 189,809,000 |
| 2020-12-31 | 4,500,000 | 2021-04-18 | 192,127,000 |
| 2021-01-09 | 9,000,000 | 2021-04-19 | 195,022,000 |
| 2021-01-13 | 10,000,000 | 2021-04-20 | 198,965,000 |
| 2021-01-26 | 22,767,000 | 2021-04-21 | 204,191,000 |
| 2021-01-31 | 24,000,000 | 2021-04-22 | 211,223,000 |
| 2021-02-03 | 31,236,000 | 2021-04-23 | 216,084,000 |
| 2021-02-09 | 40,520,000 | 2021-04-24 | 220,309,000 |
| 2021-02-28 | 52,000,000 | 2021-04-25 | 224,901,000 |
| 2021-03-14 | 64,980,000 | 2021-04-26 | 229,489,000 |
| 2021-03-20 | 74,956,000 | 2021-04-27 | 235,976,000 |
| 2021-03-22 | 80,463,000 | 2021-04-28 | 243,905,000 |
| 2021-03-23 | 82,846,000 | 2021-04-29 | 253,463,000 |
| 2021-03-24 | 85,859,700 | 2021-04-30 | 265,064,000 |
| 2021-03-25 | 91,346,000 | 2021-05-01 | 270,406,000 |
| 2021-03-26 | 97,470,000 | 2021-05-02 | 275,338,000 |
| 2021-03-27 | 102,410,000 | 2021-05-03 | 279,905,000 |
| 2021-03-28 | 106,613,000 | 2021-05-04 | 284,595,000 |
| 2021-03-29 | 110,962,000 | 2021-05-05 | 289,627,000 |
| 2021-03-30 | 114,690,000 | 2021-05-06 | 297,734,000 |

| | | | |
|---|---|---|---|
| 2021-03-31 | 119,821,000 | 2021-05-07 | 308,226,000 |
| 2021-04-01 | 126,616,000 | 2021-05-08 | 317,586,000 |
| 2021-04-02 | 133,800,999 | 2021-05-09 | 324,307,000 |
| 2021-04-03 | 136,677,000 | 2021-05-10 | 332,964,000 |
| 2021-04-04 | 139,970,000 | 2021-05-11 | 342,697,000 |
| 2021-04-05 | 142,802,000 | 2021-05-12 | 354,272,000 |
| 2021-04-06 | 145,392,000 | 2021-05-13 | 366,910,000 |
| 2021-04-07 | 149,071,000 | 2021-05-14 | 380,633,000 |
| 2021-04-08 | 155,150,000 | 2021-05-15 | 392,987,000 |
| 2021-04-09 | 161,121,000 | 2021-05-16 | 406,938,000 |
| 2021-04-10 | 164,471,000 | 2021-05-17 | 421,991,000 |
| 2021-04-11 | 167,343,000 | 2021-05-18 | 435,689,000 |
| 2021-04-12 | 171,928,000 | 2021-05-19 | 449,511,000 |
| 2021-04-13 | 175,623,000 | 2021-05-20 | 466,698,000 |
| 2021-04-14 | 179,216,000 | 2021-05-21 | 483,343,000 |
| 2021-04-15 | 183,536,000 | 2021-05-22 | 497,272,000 |
| 2021-04-16 | 187,368,000 | 2021-05-23 | 510,858,000 |

### 6 Sensitivity analyses

#### 6.1 leaky vaccine

In the baseline scenario, we showed the results considering an “all-or-nothing” vaccine model (Fig. 2). To explore the robustness of the results to the choice of the vaccine model, we used a ‘leaky’ vaccine as a sensitivity analysis. Infections of ‘leaky’ model are much bigger than ‘all-or-nothing’ model (Fig. S3).

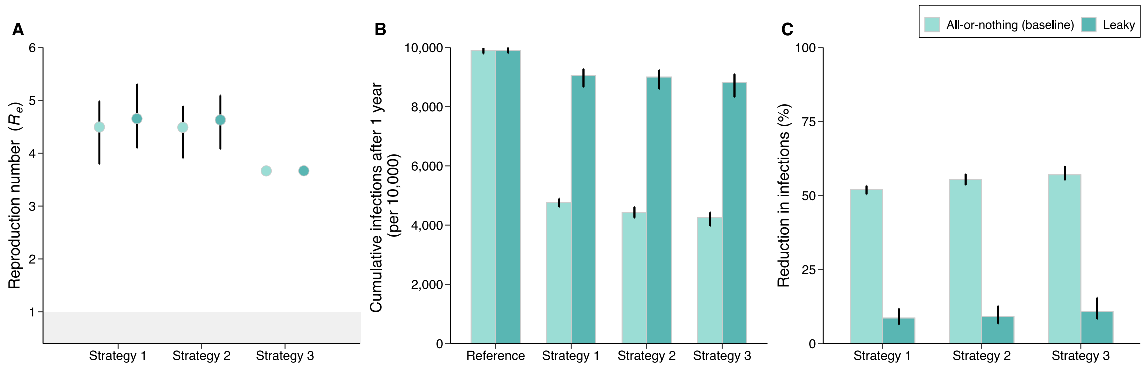

**Figure S3. Sensitivity analysis on the vaccine model. A** Estimated effective reproduction number $R_e$ (mean and 95% CI) at the start of epidemic for three analyzed vaccination strategies. **B** Cumulative number of infections (mean and 95% CI) after 1 year for the *reference scenario* and the three analyzed vaccination strategies. **C** Reduction in the number of infections (mean and 95% CI) due to vaccination with respect to the *reference scenario*.

#### 6.2 Initial number of infected individuals

In the baseline analysis, we used 40 infected individuals to initialize the epidemic (Fig. 2). To explore the robustness of the results to this choice, we varied the initial number of infected individuals by assuming 10, 20, and 100 seeds. The estimated values of $R_e$ are independent with number of seeds; the number of infections is not affected as well (Fig. S4).

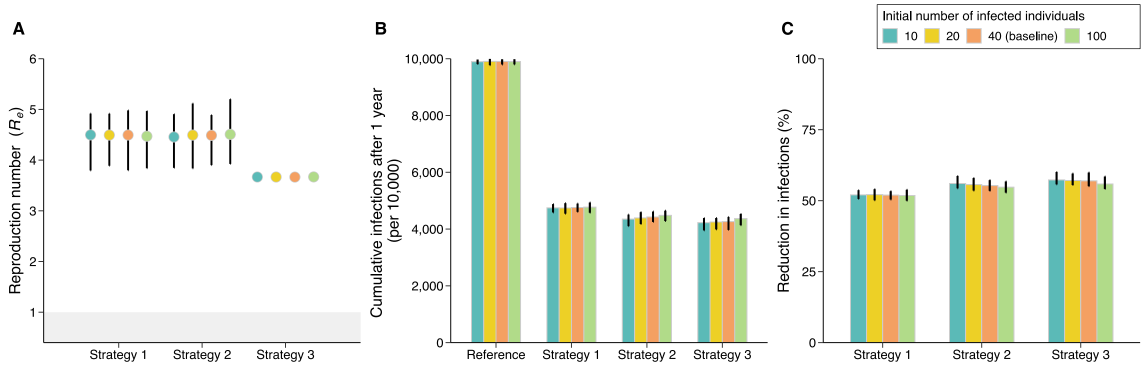

**Figure S4. Sensitivity analysis on the initial number of infected individuals. A** Estimated effective reproduction number $R_e$ (mean and 95% CI) at the start of epidemic for three analyzed vaccination strategies. **B** Cumulative number of infections (mean and 95% CI) after 1 year for the *reference scenario* and three analyzed vaccination strategies. **C** Reduction in infections (mean and 95% CI) due to vaccination with respect to the *reference scenario*.

#### 6.3 Homogeneous susceptibility to infection by age

In the main analysis, we considered age-specific susceptibility to infection [20] (Fig. 2). Here we explored a scenario where we assumed homogenous susceptibility to infection by age. All the obtained results are very consistent with those obtained in the main analysis (Fig. S5).

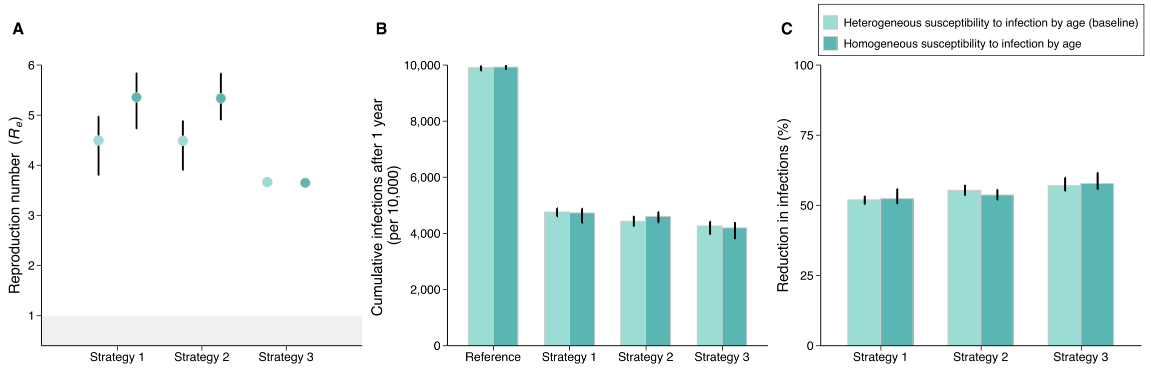

**Figure S5. Sensitivity analysis assuming homogeneous susceptibility to infection by age. A** Estimated effective reproduction number $R_e$ (mean and 95% CI) at the start of epidemic for the three analyzed vaccination strategies. **B** Cumulative number of infections (mean and 95% CI) after 1 year for the *reference scenario* and the three analyzed vaccination strategies. **C** Reduction in infections (mean and 95% CI) due to vaccination with respect to the *reference scenario*.

#### 6.4 Natural immunity

In the baseline analysis, we considered a population with no pre-existing natural immunity (Fig. 2). We performed a sensitivity analysis where we considered that 10%, 20%, and 30% of the population had natural immunity before the start of the epidemic. The addition of natural immunity remarkably affects the results both in terms of transmissibility potential (Fig. S6A) and infections (Fig. S6B-C).

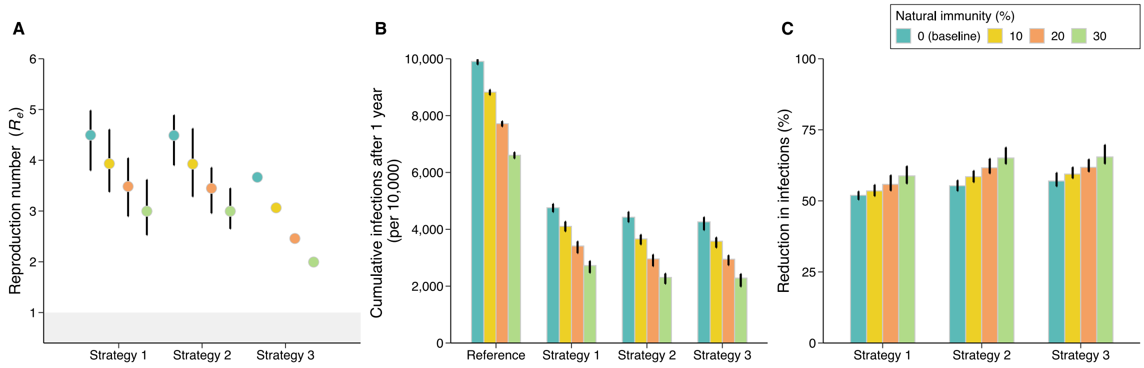

**Figure S6. Sensitivity analysis on the natural immunity. A** Estimated effective reproduction number $R_e$ (mean and 95% CI) at the start of epidemic for the three analyzed vaccination strategies. **B** Cumulative number of infections (mean and 95% CI) after 1 year for the *reference scenario* and the three analyzed vaccination strategies. **C** Reduction in infections (mean and 95% CI) due to vaccination with respect to the *reference scenario*.

##### 6.5 Vaccine efficacy

In the baseline analysis, we considered a two-dose vaccine whose efficacy against SARS-CoV-2 infections was set to 54.3% [21-23] for all age groups (Fig. 2). Here we performed three sensitivity analyses on the VE. In the first one, we varied the overall vaccine efficacy up to 79%, in line with the vaccine effectiveness for BioNTech/Pfizer [34] (Fig. S7); second, we varied the vaccine efficacy for individuals aged 3-17 and 60+ years relative to that of individuals aged 18-59 years (Fig. S8); third, we use explored different times from the administration of the second dose and maximum protection (Fig. S9).

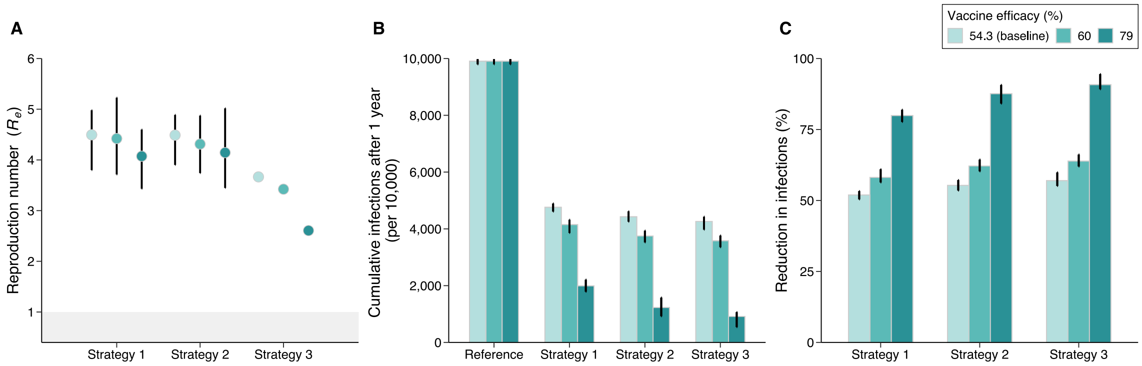

**Figure S7. Sensitivity analysis on the vaccine efficacy. A** Estimated effective reproduction number $R_e$ (mean and 95% CI) at the start of epidemic for the three analyzed vaccination strategies. **B** Cumulative number of infections (mean and 95% CI) after 1 year for the *reference scenario* and the three analyzed vaccination strategies. **C** Reduction in infections (mean and 95% CI) due to vaccination with respect to the *reference scenario*.

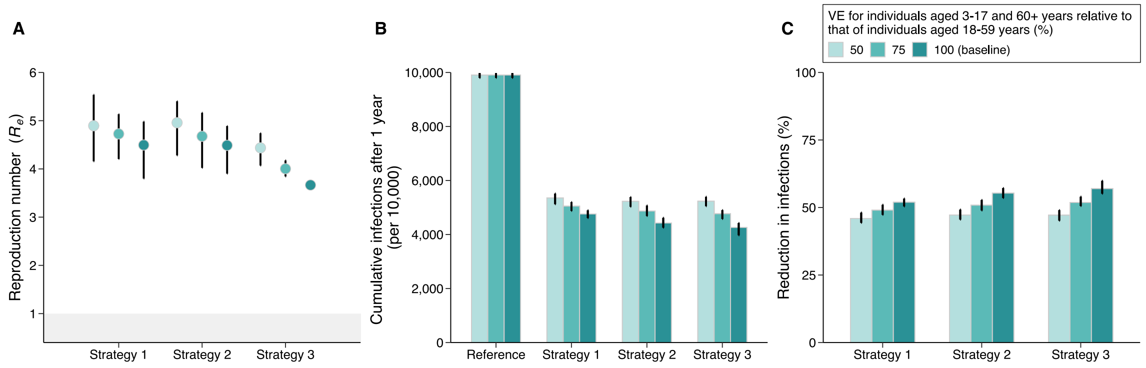

**Figure S8. Sensitivity analysis on the vaccine efficacy for individuals aged 3-17 and 60+ years relative to that of individuals aged 18-59 years.** **A** Estimated reproduction number $R_e$ (mean and 95% CI) at the start of epidemic for the three analyzed vaccination strategies. **B** Cumulative number of infections (mean and 95% CI) after 1 year for the *reference scenario* and the three analyzed vaccination strategies. **C** Reduction in the number of infections (mean and 95% CI) due to vaccination with respect to the *reference scenario*.

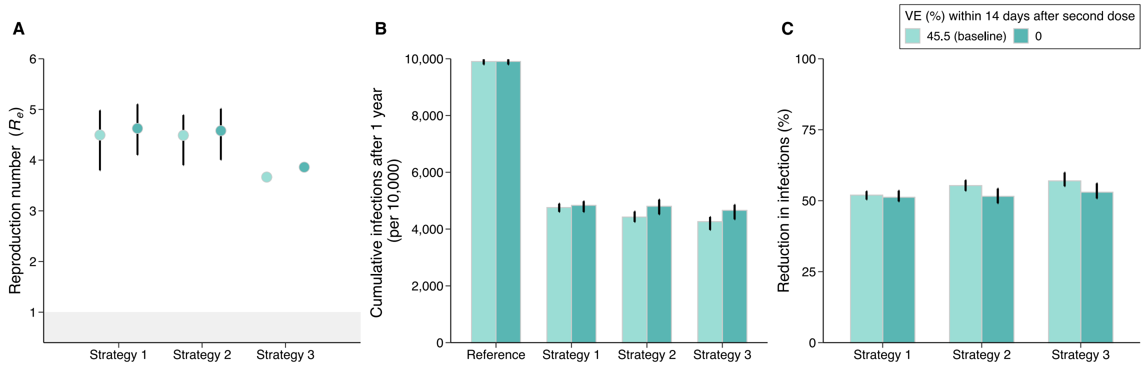

**Figure S9. Sensitivity analysis of the time between vaccination and maximum protection.** **A** Estimated effective reproduction number $R_e$ (mean and 95% CI) at the start of epidemic for the three analyzed vaccination strategies. **B** Cumulative number of infections (mean and 95% CI) after 1 year for the *reference scenario* and the three analyzed vaccination strategies. **C** Reduction in the number of infections (mean and 95% CI) due to vaccination with respect to the *reference scenario*.

#### 6.6 Time interval between the two doses

In the baseline analysis, we considered a two-dose vaccine with a 21-day interval between the two doses (Fig. 2). Here we proposed a sensitivity analysis on this time interval, assuming a 14-day interval. All the obtained results are very consistent with those obtained in the baseline analysis (Fig. S10).

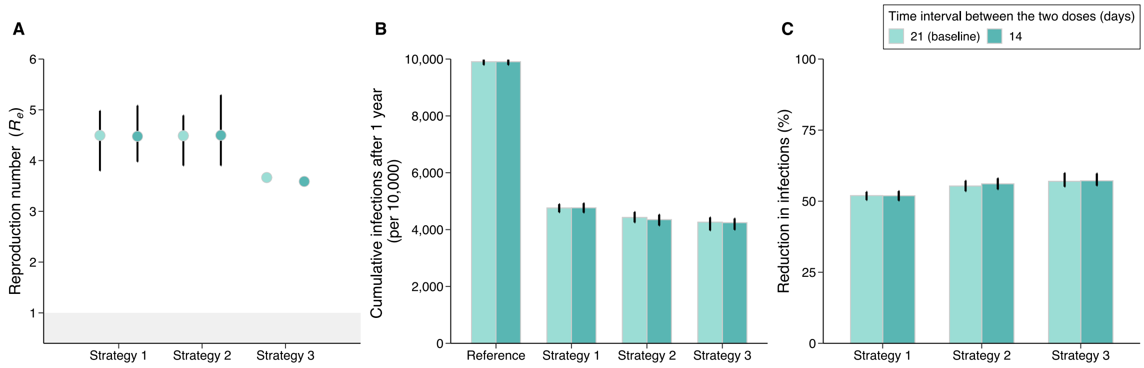

**Figure S10. Sensitivity analysis on time intervals between the two doses. A** Estimated effective reproduction number $R_e$ (mean and 95% CI) at the start of epidemic for the three analyzed vaccination strategies. **B** Cumulative number of infections (mean and 95% CI) after 1 year for the *reference scenario* and the three analyzed vaccination strategies. **C** Reduction in the number of infections (mean and 95% CI) due to vaccination with respect to the *reference scenario*.

#### 7 Scenario 1: Delaying the start of the epidemic

We tested to what extent the start of a new epidemic wave needs to be delayed (e.g., by keeping strict restriction for international travels) to allow the immunity to build up in the population, potentially reaching herd immunity levels in Fig. 3A-C. Here we reported the impact of delaying the start of the epidemic on vaccine coverage (Fig. S11A-C) and daily incidence of new infections (Fig. S11D-F).

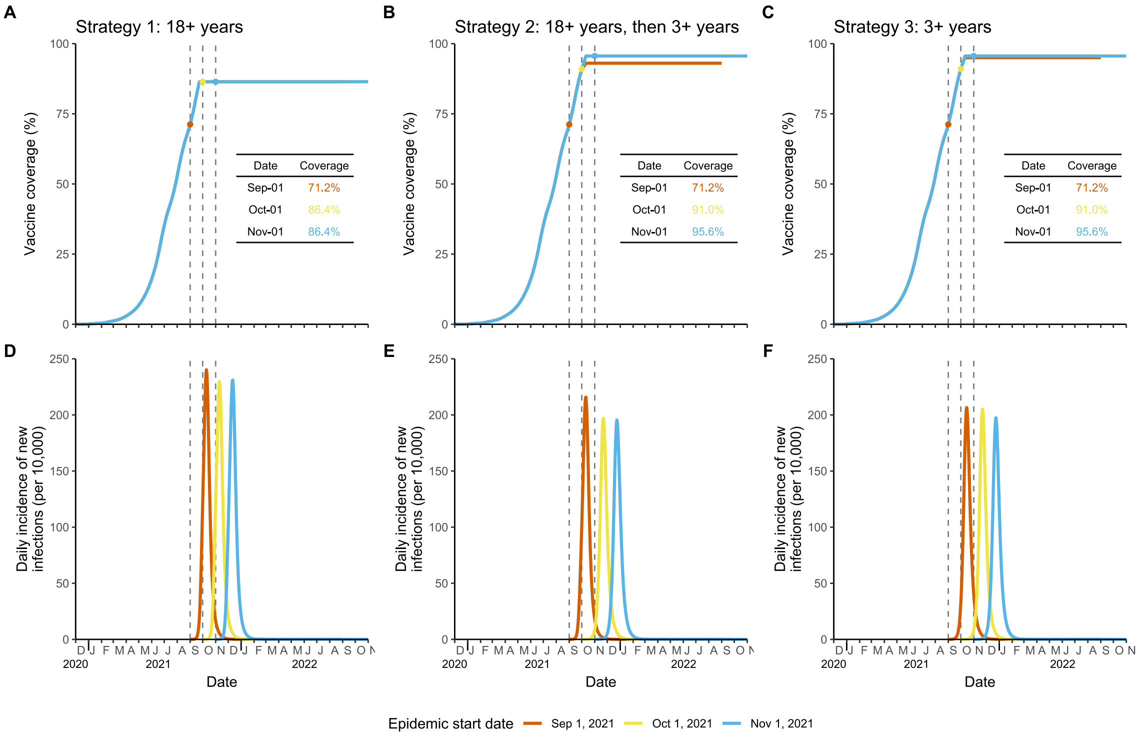

**Figure S11. Impact of delaying the start of the epidemic on vaccine coverage and daily incidence of new infections. A** Cumulative vaccination coverage of strategy 1. **B** As A, but for strategy 2. **C** As A, but for strategy 3. **D** Daily incidence of new infection for strategy 1. **E** As A, but for strategy 2. **F** As A, but for strategy 3.

#### 8 Scenario 2: Adopting NPIs in case of a new outbreak

We investigated the synergetic effect of vaccination programs combined with non-pharmaceutical interventions (NPIs) of different intensity in Fig. 3D-F. Here we showed the results of lager values of $R_0^{NPIs}$ (Fig. S12) and reported the impact of adopting NPIs on daily incidence of new infections (Fig. S13).

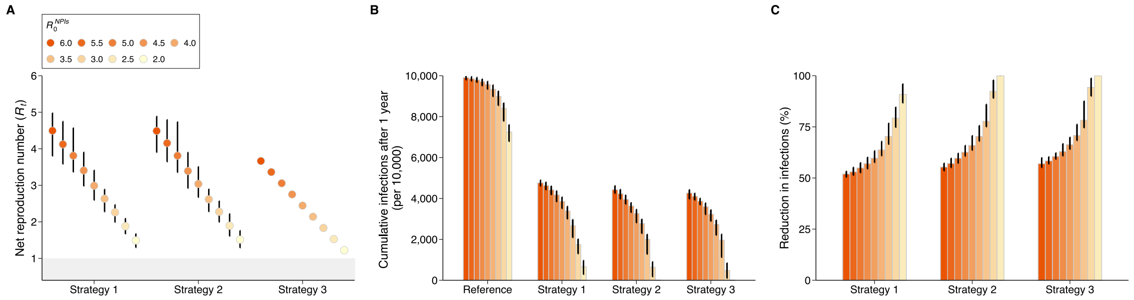

**Figure S12. Impact of adopting NPIs in case of a new outbreak. A** estimated net reproduction number ($R_t$, mean and 95% CI) adopting different intensity of NPIs, $R_0^{NPIs}$. **B** Cumulative number of infections per 10,000 individuals after 1 simulated year for *reference scenario* and three vaccination strategies (mean and 95% CI). *Reference scenario* indicates no vaccination and no NPIs with $R_0$=6.0 at the beginning of transmission. **C** Reduction in infections (mean and 95% CI) with respect to the *reference scenario*.

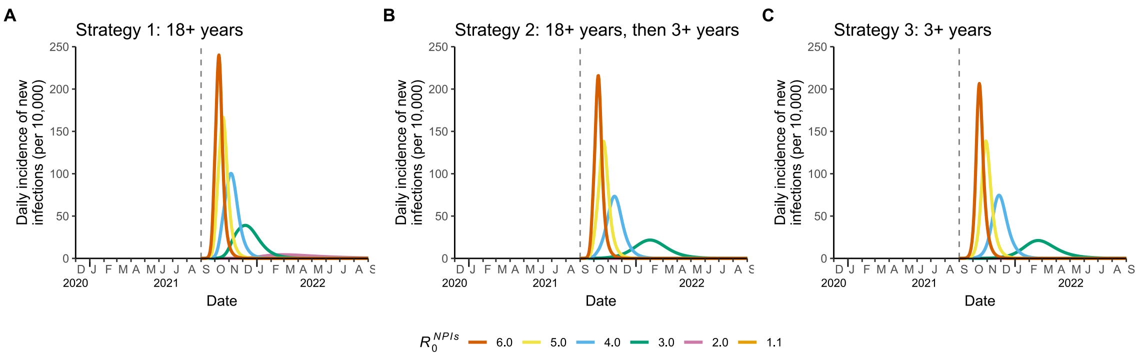

**Figure S13. Impact of adopting NPIs in case of a new outbreak on daily incidence of new infections. A** Vaccine coverage of strategy 1. **B** Vaccine coverage of strategy 2. **C** Vaccine coverage of strategy 3.

#### 9 Scenario 3: Delaying the start of the epidemic and adopting NPIs

To further improve the potential for vaccination-induced herd immunity against the SARS-CoV-2 Delta variant, we tested the combination of two scenarios mentioned above: delaying the start of the epidemic and adopting NPIs in case of a new outbreak in Fig. 4. Here we reported the impact of the above two scenarios on infections. In particular, the reduction in infections is more than 50% (Fig. S14). Besides, we performed other sensitivity analyses: exploring impact of the contact matrix considering an independently estimated contact matrix for the Chinese population [35] (Fig. S15). The obtained results show that $R_e$ is very consistent with those obtained in the main analysis (Fig. S16).

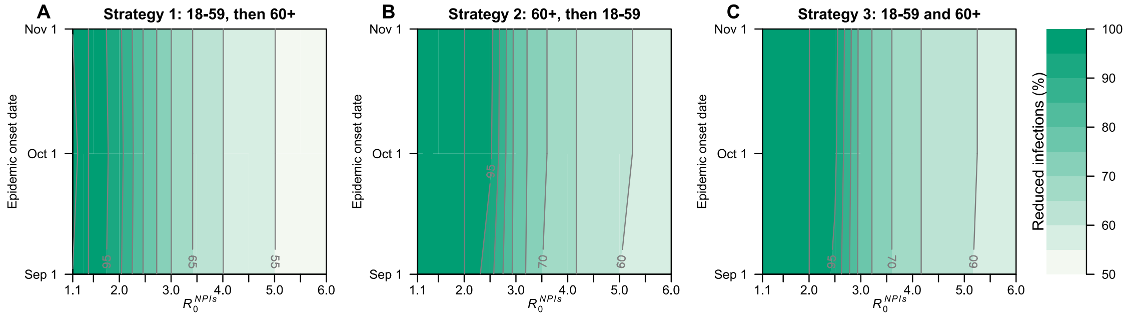

**Figure S14. Impact of delaying the start of the epidemic, and adopting NPIs on infections. A** Reduction in infections due to vaccination with respect to the *reference scenario* for strategy 1. **B** As A, but for strategy 2. **C** As A, but for strategy 3.

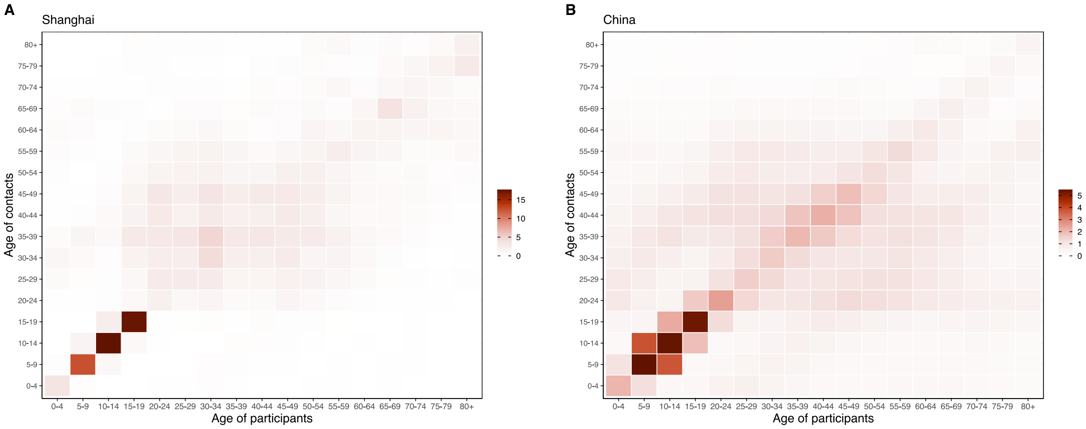

**Figure S15. Comparison of contact matrix in Shanghai and China. A** Shanghai contact matrix. **B** Chinese contact matrix [35].

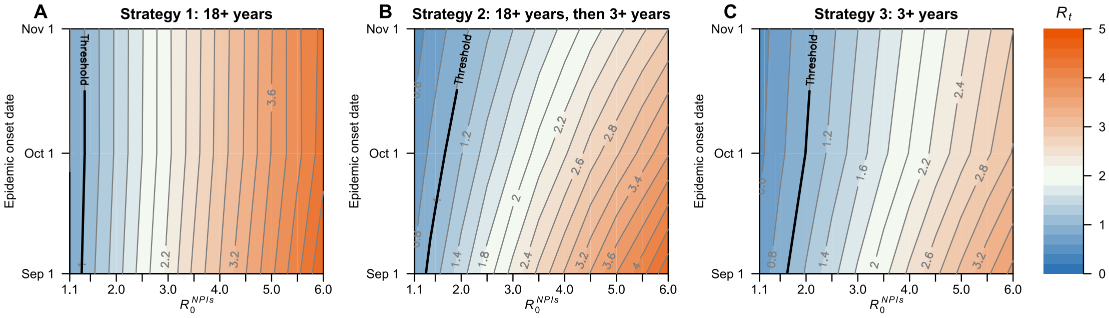

**Figure S16. Impact of delaying the start of the epidemic start and adopting NPIs on estimated net reproduction number $R_t$ using China contact matrix [35]. A** Estimated net reproduction number ($R_t$) as a function of $R_0^{NPIs}$ and epidemic start date for strategy 1. The bold line in black indicates the herd immunity threshold $R_t$ =1. **B** As A, but for strategy 2. **C** As A, but for strategy 3.

#### 10 Additional results about the herd immunity threshold

We showed the relation between $R_e$ and vaccine coverage using an 95% efficacious vaccine (Fig. S17). The obtained results show that the herd immunity against Delta strain can be reached by using a higher efficacy and level of immunity needed to lead the effective reproduction number below the epidemic threshold is lower if vaccination is extended to individuals aged 3-17 years early on. Besides, we showed the impact of vaccine efficacy and vaccine coverage on estimated net reproduction number $R_t$ in Fig. 5 without adopting NPIs. Here we performed sensitivity analysis: exploring impact of adopting different intensity of NPIs (Fig. S18). The obtained results show that the net reproduction number can be reduced below the unit in the presence of NPIs.

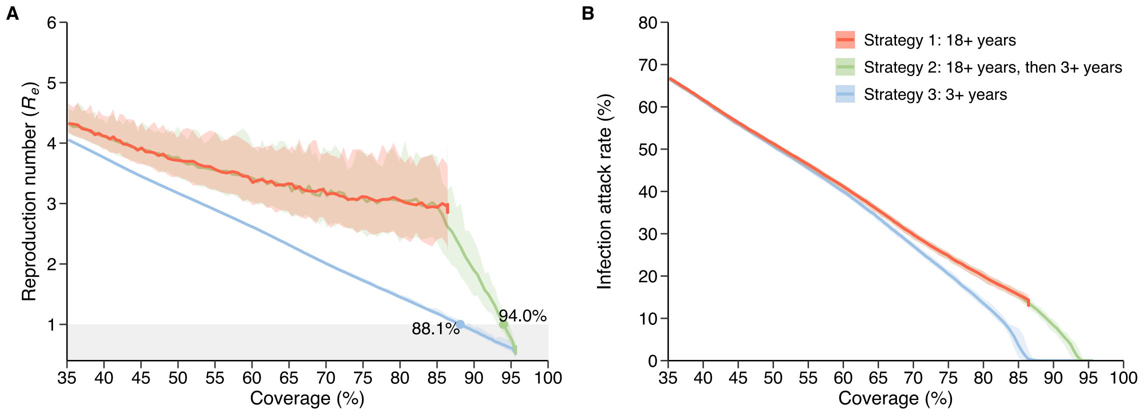

**Figure S17. Impact of vaccine efficacy on the reproduction number $R_e$ and the infection attack rate.** A Estimated effective reproduction number $R_e$ (mean and 95% CI) as a function of the vaccination coverage, assuming an 95% efficacious vaccine and no transmission. Colors refer to the three vaccination strategies. The shaded area in gray indicates the herd immunity threshold $R_e$=1. The numbers around the line indicate where $R_e$ crosses this threshold. B The infection attack rate after 1 simulated year as a function of vaccine coverage with an 95% efficacious vaccine. The transmission is simulated with the initial vaccine coverage and there is no longer vaccination after the infection is seeded.

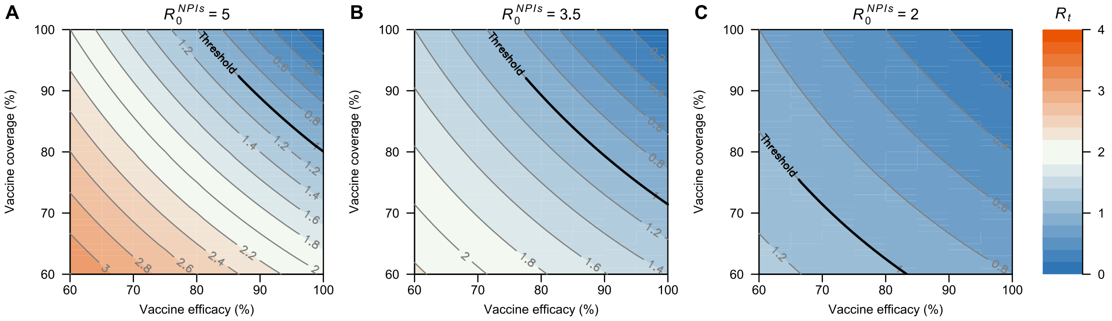

**Figure S18. Impact of vaccine efficacy and vaccine coverage on estimated net reproduction umber $R_t$ under different intensity of NPIs. A** $R_0^{NPIs} = 5.0$. **B** $R_0^{NPIs} = 3.5$. **C** $R_0^{NPIs} = 2.0$.d

#### 11 US-like scenario

In the baseline analysis, we considered the scenario in China with VE=54.3% [21-23] and no initial natural immunity [19] (Fig. 2). Here we proposed a sensitivity analysis where we simulated a US-like scenario, where VE=79% [34] and the initial fraction of naturally immune population is set at 22% [36], finding remarkably different results both in terms of transmission potential and number of infections (Fig. S19).

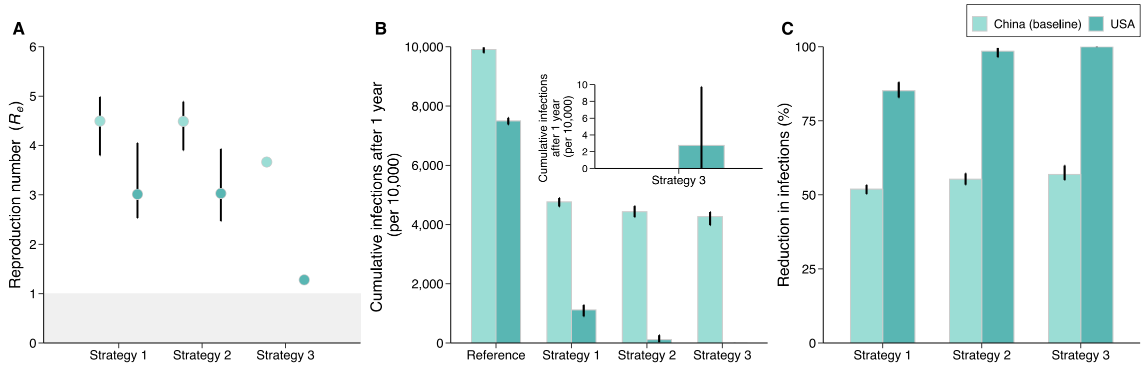

**Figure S19. Comparison between a US-like scenario (VE=79% [34], initial fraction of naturally immune population=22% [36]) and China (VE=54.3% [21], no initial natural immunity [19]). A** Estimated reproduction number $R_e$ (mean and 95% CI) at the start of epidemic for the three analyzed vaccination strategies. **B** Cumulative number of infections (mean and 95% CI) after 1 year for the *reference scenario* and the three analyzed vaccination strategies. **C** Reduction in the number of infections (mean and 95% CI) due to vaccination with respect to the *reference scenario*.
